# Brain structure and function show distinct relations with genetic predispositions to mental health and cognition

**DOI:** 10.1101/2021.03.07.21252728

**Authors:** Shu Liu, Dirk J.A. Smit, Abdel Abdellaoui, Guido A. van Wingen, Karin J.H. Verweij

## Abstract

Mental health and cognitive achievement are partly heritable. To identify the underlying neural mechanisms, we associated genetic predispositions to various mental health and cognitive traits with a large set of structural and functional brain measures from the UK Biobank (N=36,799). We show that genetic predispositions to attention deficit hyperactivity disorder, smoking initiation, and cognitive traits have stronger associations with brain structure than with brain function, whereas genetic predispositions to most other psychiatric disorders have stronger associations with brain function than with brain structure. These results reveal that genetic predispositions to mental health and cognitive traits have distinct brain profiles.

## Introduction

Twin and family studies have revealed that mental health and cognitive traits are partly heritable^1^. Genome-wide association studies (GWAS) have identified many single-nucleotide polymorphisms (SNPs) across the whole genome associated with these traits^2^. Each trait is influenced by many genes that individually explain a very small part of the variance. This polygenic nature poses a challenge to understanding the genetic architecture and biological mechanisms underlying specific traits^3^. A promising approach that we will explore here is the use of imaging genetics to examine brain-based intermediate phenotypes, also known as endophenotypes^4, 5^. The effects of genes are not expressed directly at the level of behavior, but are more likely mediated by their molecular and cellular effects on brain development and information processing^6, 7^. The neural mechanisms through which genetic variants influence one’s cognitive capacity and mental health remain largely unknown.

The cerebral cortex is the outer layer of the brain that contains the neuronal cell bodies, also known as gray matter. It forms a folded sheet that plays a major role in multiple aspects of higher cognitive function. A recent study reported that variation in total cortical surface area (CSA) was genetically correlated with cognitive function, depression, and attention deficit hyperactivity disorder (ADHD), but not with many other psychiatric disorders^8^. Another study found significant genetic associations between total brain volume and cognitive traits including general cognitive function, educational outcomes, intelligence, and numerical reasoning^9^. Moreover, genetic predisposition to cognition has been consistently associated with brain structure^10, 11^, but few studies focused on the genetic relationship between cognition and brain function^12^. In contrast, psychiatric disorders, especially schizophrenia and bipolar disorder, have been found to be genetically associated with variation in brain function^13–15^, but inconsistent results regarding the association between polygenic risk for psychiatric disorders and brain structure were observed in previous studies^16–18^. While studies combining genetic and neuroimaging data have made considerable progress in revealing neural correlates of various traits, there are still some limitations: (1) the sample sizes of most studies have been relatively limited, resulting in insufficient statistical power; (2) most studies only focused on one specific trait, which makes it difficult to compare the association patterns between different traits; (3) few studies have combined structural and functional neuroimaging information, leaving open whether certain predispositions are predominantly linked to brain structure or function.

In the present study, we systematically investigated the genetic associations of multiple mental health and cognitive traits with structural and functional neuroimaging measures. We used both polygenic score (PGS)^19^ analyses and genome-wide genetic correlations based on GWAS summary statistics^20^. We calculated PGSs for 14 mental health and cognitive traits which represent the aggregated genetic effects across the genome, and assessed their associations with structural and functional neuroimaging measures. In addition, we also estimated genetic correlations of the mental health and cognitive traits with neuroimaging features based on genome-wide single-nucleotide polymorphism (SNP) data using linkage disequilibrium score (LDSC) regression. Genetic correlations can provide a good indication of the magnitude of the overlap in genetic influences, but they are less powerful than PGSs to detect significant associations. Finally, we modeled the joint genetic associations of the mental health and cognitive traits with brain structure and function, using a robust partial least squares (PLS) regression^21^. To decompose the association patterns of various traits with brain structure and function, PLS regression is used to identify a limited number of latent variables that link the many genotype variables to the many imaging phenotypes.

## RESULTS

We analyzed genotypic and structural and functional neuroimaging data from the UK Biobank^22^ (N = 36,799). Structural measures from T1-weighted imaging included 3 global measures (intra-cranial volume (ICV), total CSA, and average cortical thickness (CT)) and 146 regionally cortical and subcortical measures. Functional brain measures included the activity of 21 resting-state networks and 210 functional connectivities (FCs) between them. We used the genotype data (∼1.3 million SNPs) to build PGSs for seven mental health disorders (ADHD, Alzheimer’s disease, autism spectrum disorder (ASD), bipolar disorder, eating disorder, major depression, schizophrenia), five substance use traits (cigarettes per day, smoking initiation, smoking cessation, caffeine consumption, alcoholic drinks per week), and two cognitive traits (cognitive ability and educational attainment). Then, we investigated the associations between PGSs for these 14 mental health and cognitive traits and the structural and functional brain measures.

In addition, to quantify overlapping genetic effects, we also performed LDSC regression analyses to estimate genetic correlations of these traits with the brain measures using GWAS summary statistics. The summary statistics for the 14 mental health and cognitive traits were obtained from existing GWASs (**Supplementary Table 1**). For brain structural and functional measures, we conducted the GWASs in the UK Biobank sample. SNP-based heritability estimates as obtained from LDSC for the brain measures can be found in Supplementary Fig. 1 and **Supplementary Table 2**. SNP-based heritability estimates ranged between 0 and 37.8%, with higher estimated for total CSA and ICV, and lower estimates for the FCs.

### Genetic relationships with global brain structure

Table 1 shows the associations of PGSs for the 14 mental health and cognitive traits with three global structural measures, accounting for the effects of age, sex, MRI scanning positions, and the first 25 genetic principal components. To account for three sets of analyses (global structural measures, regional structural measures, and functional measures), we applied a false discovery rate (FDR) multiple testing correction with a significance threshold of 0.0167 (0.05/3) (see **Methods**).

**Table 1.**
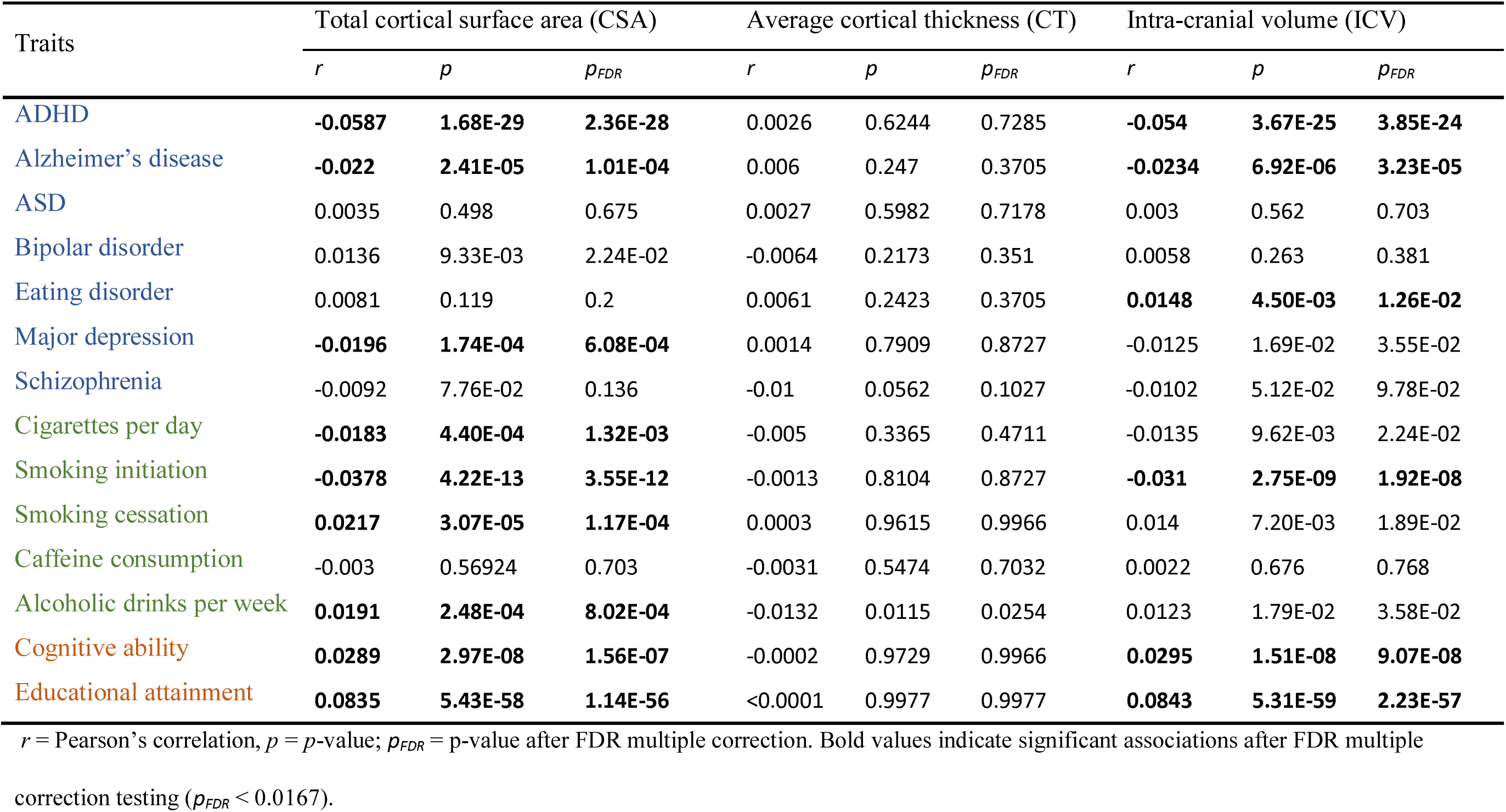
Correlations of 14 PGSs for mental health and cognitive traits with three global structural brain measures.

| Traits | Total cortical surface area (CSA) |  |  | Average cortical thickness (CT) |  |  | Intra-cranial volume (ICV) |  |  |
| --- | --- | --- | --- | --- | --- | --- | --- | --- | --- |
|  | *r* | *p* | *p<sub>FDR</sub>* | *r* | *p* | *p<sub>FDR</sub>* | *r* | *p* | *p<sub>FDR</sub>* |
| ADHD | **-0.0587** | **1.68E-29** | **2.36E-28** | 0.0026 | 0.6244 | 0.7285 | **-0.054** | **3.67E-25** | **3.85E-24** |
| Alzheimer's disease | **-0.022** | **2.41E-05** | **1.01E-04** | 0.006 | 0.247 | 0.3705 | **-0.0234** | **6.92E-06** | **3.23E-05** |
| ASD | 0.0035 | 0.498 | 0.675 | 0.0027 | 0.5982 | 0.7178 | 0.003 | 0.562 | 0.703 |
| Bipolar disorder | 0.0136 | 9.33E-03 | 2.24E-02 | -0.0064 | 0.2173 | 0.351 | 0.0058 | 0.263 | 0.381 |
| Eating disorder | 0.0081 | 0.119 | 0.2 | 0.0061 | 0.2423 | 0.3705 | **0.0148** | **4.50E-03** | **1.26E-02** |
| Major depression | **-0.0196** | **1.74E-04** | **6.08E-04** | 0.0014 | 0.7909 | 0.8727 | -0.0125 | 1.69E-02 | 3.55E-02 |
| Schizophrenia | -0.0092 | 7.76E-02 | 0.136 | -0.01 | 0.0562 | 0.1027 | -0.0102 | 5.12E-02 | 9.78E-02 |
| Cigarettes per day | **-0.0183** | **4.40E-04** | **1.32E-03** | -0.005 | 0.3365 | 0.4711 | -0.0135 | 9.62E-03 | 2.24E-02 |
| Smoking initiation | **-0.0378** | **4.22E-13** | **3.55E-12** | -0.0013 | 0.8104 | 0.8727 | **-0.031** | **2.75E-09** | **1.92E-08** |
| Smoking cessation | **0.0217** | **3.07E-05** | **1.17E-04** | 0.0003 | 0.9615 | 0.9966 | 0.014 | 7.20E-03 | 1.89E-02 |
| Caffeine consumption | -0.003 | 0.56924 | 0.703 | -0.0031 | 0.5474 | 0.7032 | 0.0022 | 0.676 | 0.768 |
| Alcoholic drinks per week | **0.0191** | **2.48E-04** | **8.02E-04** | -0.0132 | 0.0115 | 0.0254 | 0.0123 | 1.79E-02 | 3.58E-02 |
| Cognitive ability | **0.0289** | **2.97E-08** | **1.56E-07** | -0.0002 | 0.9729 | 0.9966 | **0.0295** | **1.51E-08** | **9.07E-08** |
| Educational attainment | **0.0835** | **5.43E-58** | **1.14E-56** | <0.0001 | 0.9977 | 0.9977 | **0.0843** | **5.31E-59** | **2.23E-57** |

*r* = Pearson's correlation, *p* = *p*-value; *p<sub>FDR</sub>* = *p*-value after FDR multiple correction. Bold values indicate significant associations after FDR multiple
correction testing (*p<sub>FDR</sub>* < 0.0167).

The PGSs showed many significant associations with total CSA and ICV, but none with average CT. Higher polygenic risks for ADHD, Alzheimer’s disease, major depression, cigarettes per day, and smoking initiation were strongly associated with *lower* total CSA. Conversely, higher PGSs for smoking cessation, alcoholic drinks per week, cognitive ability, and educational attainment were strongly associated with *higher* total CSA. Similarly, higher polygenic risks for ADHD, smoking initiation, and Alzheimer’s disease were significantly associated with *lower* ICV, whereas higher PGSs for cognitive ability, educational attainment, and eating disorder were significantly associated with *higher* ICV. Polygenic risks for ASD, bipolar disorder, schizophrenia and caffeine consumption were not significantly associated with any global measure of brain structure.

The results of the genetic correlation analysis for global structural measures were largely in line with those of the PGS analysis (**Supplementary Table 3**). Total CSA and ICV showed highly significant genetic correlations with ADHD (*r_g_* = −0.170 and *r_g_* = −0.159), cognitive ability (*r_g_* = 0.230 and *r_g_* = 0.228) and educational attainment (*r_g_* = 0.211 and *r_g_* = 0.159), but no significant correlations were observed with average CT. Most psychiatric disorders (ASD, bipolar disorder, eating disorder, schizophrenia) were not genetically correlated with measures of global brain structure.

### Genetic relationships with regional brain structure

We investigated the associations between the 14 PGSs and the volumes of 14 subcortical regions, as well as CSA and CT of 66 cortical regions based on the Desikan-Killiany (DK) atlas. These analyses were corrected for ICV, total CSA, or average CT, respectively, to avoid overlap with the analysis of global brain structure and identify regional specificity of the relationships.

We found six significant associations for subcortical volumes (Fig. 1, **Supplementary Table 4**). Notably, volume of the thalamus was negatively associated with genetic vulnerability to schizophrenia and positively with genetic predisposition to educational attainment. In addition, the PGS for major depression was positively associated with volume of the left caudate nucleus, the PGS for Alzheimer’s disease was negatively associated with volume of the left putamen, and the PGS for cigarettes per day showed negative associations with volume of the right putamen. As for genetic correlation analysis, we only found a significant correlation between volume of the thalamus and schizophrenia (*r_g_* = −0.135) (Supplementary Fig. 2, **Supplementary Table 5**).

**Fig 1.**
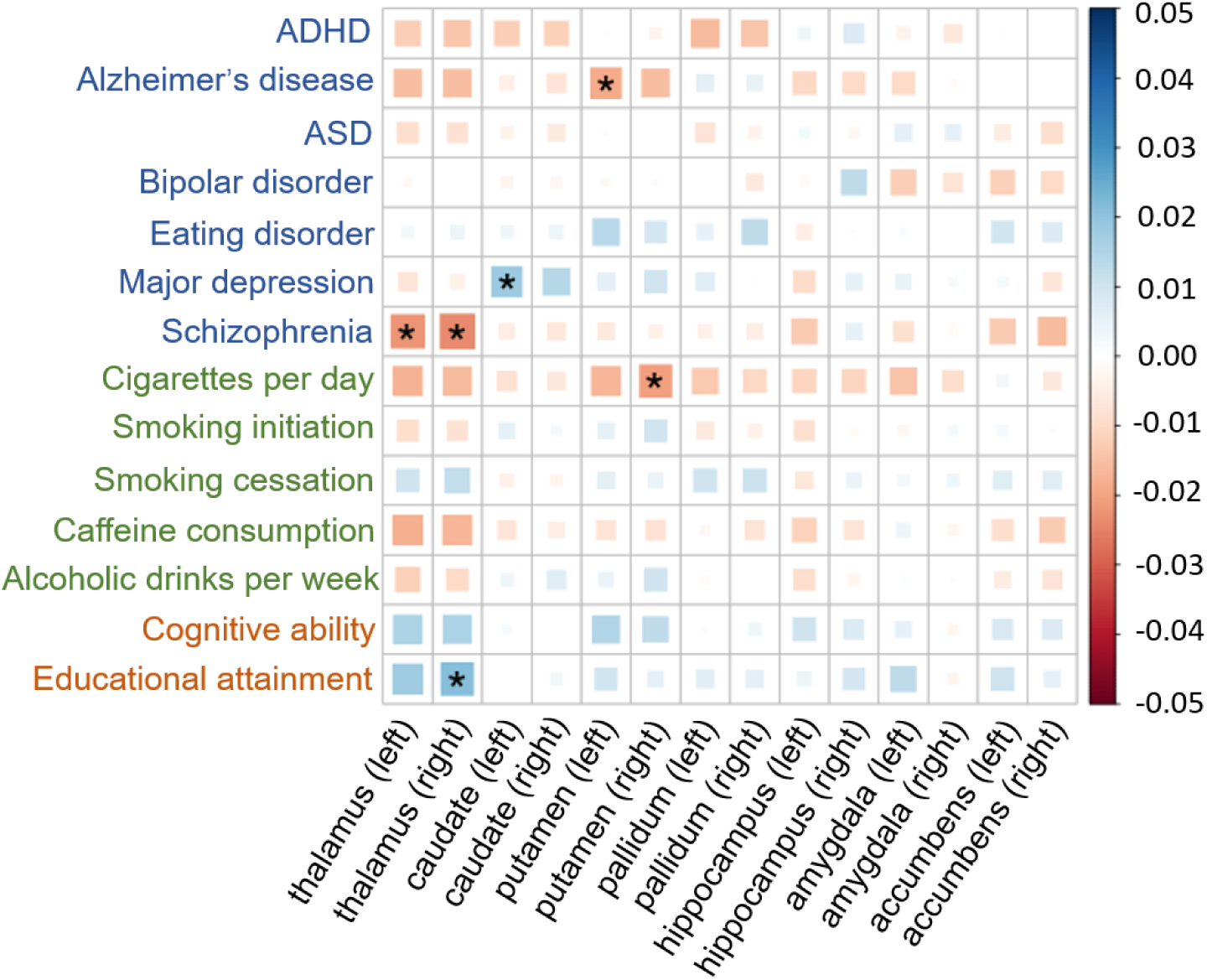
Associations between the 14 PGSs and the 14 subcortical volumes, corrected for ICV. The size and color of each box indicate the magnitude of the association. Blue squares indicate positive associations, and red squares indicate negative associations. Asterisks indicate significant associations after FDR multiple testing correction.

For CSA of brain regions, a total of 27 significant associations were found with the 14 PGSs (Fig. 2, **Supplementary Table 6**). Of them, the PGS for educational attainment was mainly positively associated with CSA in regions of the inferior frontal cortex and negatively associated with CSA in regions of occipital cortex. Genetic vulnerability to cognitive ability was also found to be associated with higher CSA of some frontal regions. Interestingly, PGSs for ASD, bipolar disorder, and major depression all showed significant positive associations with CSA of the cingulate regions. In addition, the PGS for smoking initiation showed significant negative associations with CSA of frontal regions, whereas the PGS for alcoholic drinks per week was negatively associated with CSA of medial inferior occipital cortex. The genetic correlation analysis found largely comparable results though with fewer significant associations, with significant genetic correlations of cognitive ability and educational attainment with CSA of the pars orbitalis of the inferior frontal gyrus (*r*_g_ = 0.147 and *r*_g_ = 0.170) (Supplementary Fig. 3, **Supplementary Table 7**).

**Fig 2.**
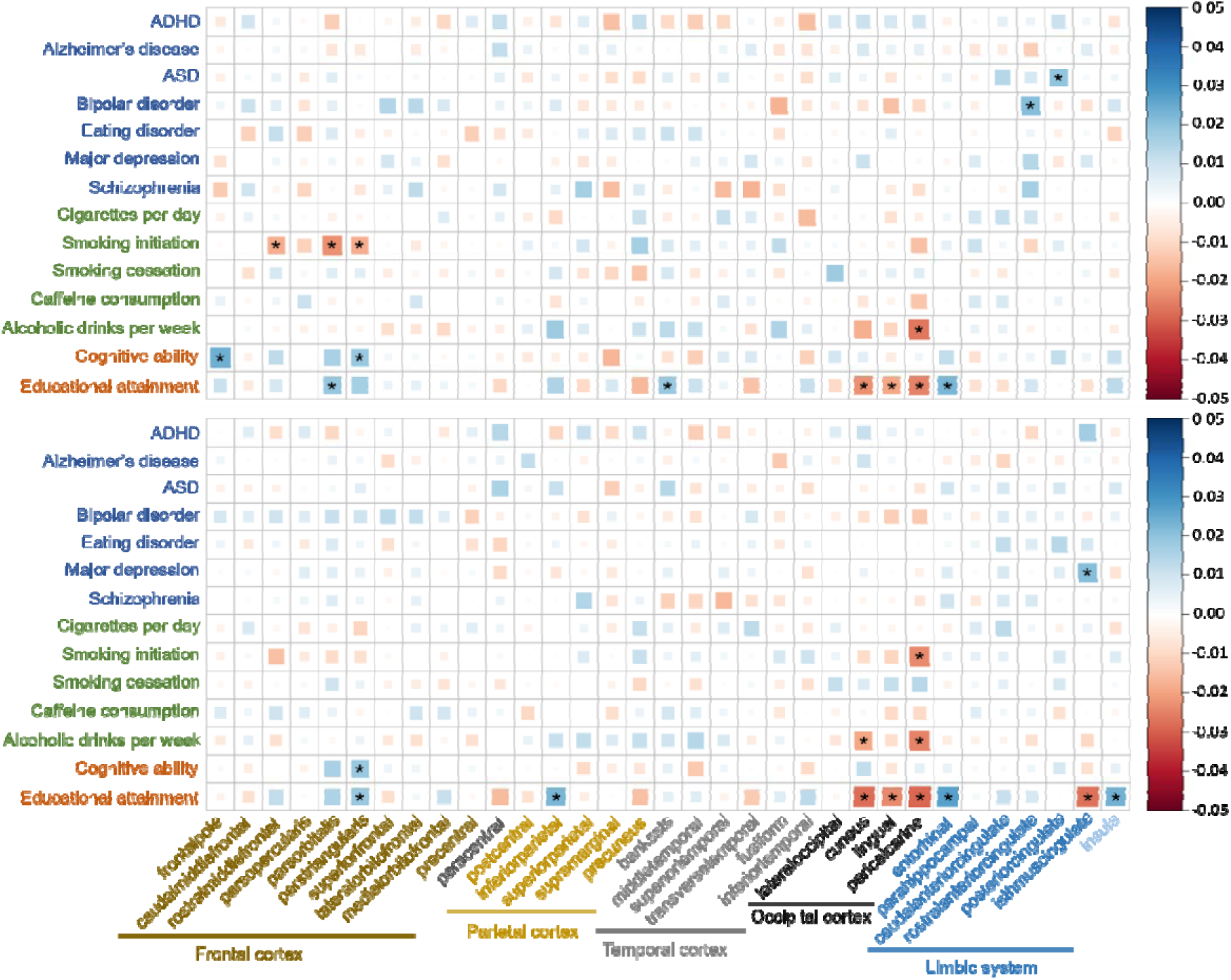
Associations between the 14 PGSs and the CSA of 66 cortical regions, corrected for total CSA. The size and color of each box indicate the magnitude of the association. Blue squares indicate positive associations, and red squares indicate negative associations. The upper half shows the PGS-CSA correlation map across the left hemisphere, and the lower half across right hemisphere. Asterisks indicate significant associations after FDR multiple testing correction.

Although the global measure of CT did not show any significant associations with PGSs, we found 27 significant associations with regional CT (Fig. 3, **Supplementary Table 8**). For cognitive traits, the PGS for educational attainment showed widespread significant negative associations with CT of some parietal regions and positive associations with CT of the superior temporal cortex. Genetic predisposition to higher cognitive ability was also significantly associated with higher CT of the superior temporal cortex. For mental health disorders, CT of the orbitofrontal cortex was found to be negatively associated with PGSs for bipolar disorder and schizophrenia. For substance use traits, CT of the superior frontal cortex was significantly associated with PGSs for smoking cessation and alcoholic drinks per week. The genetic correlation analysis showed a negative correlation of educational attainment with CT of the inferior parietal cortex and two positive correlations with CT of the superior temporal regions. (Supplementary Fig. 4, **Supplementary Table 9**).

**Fig 3.**
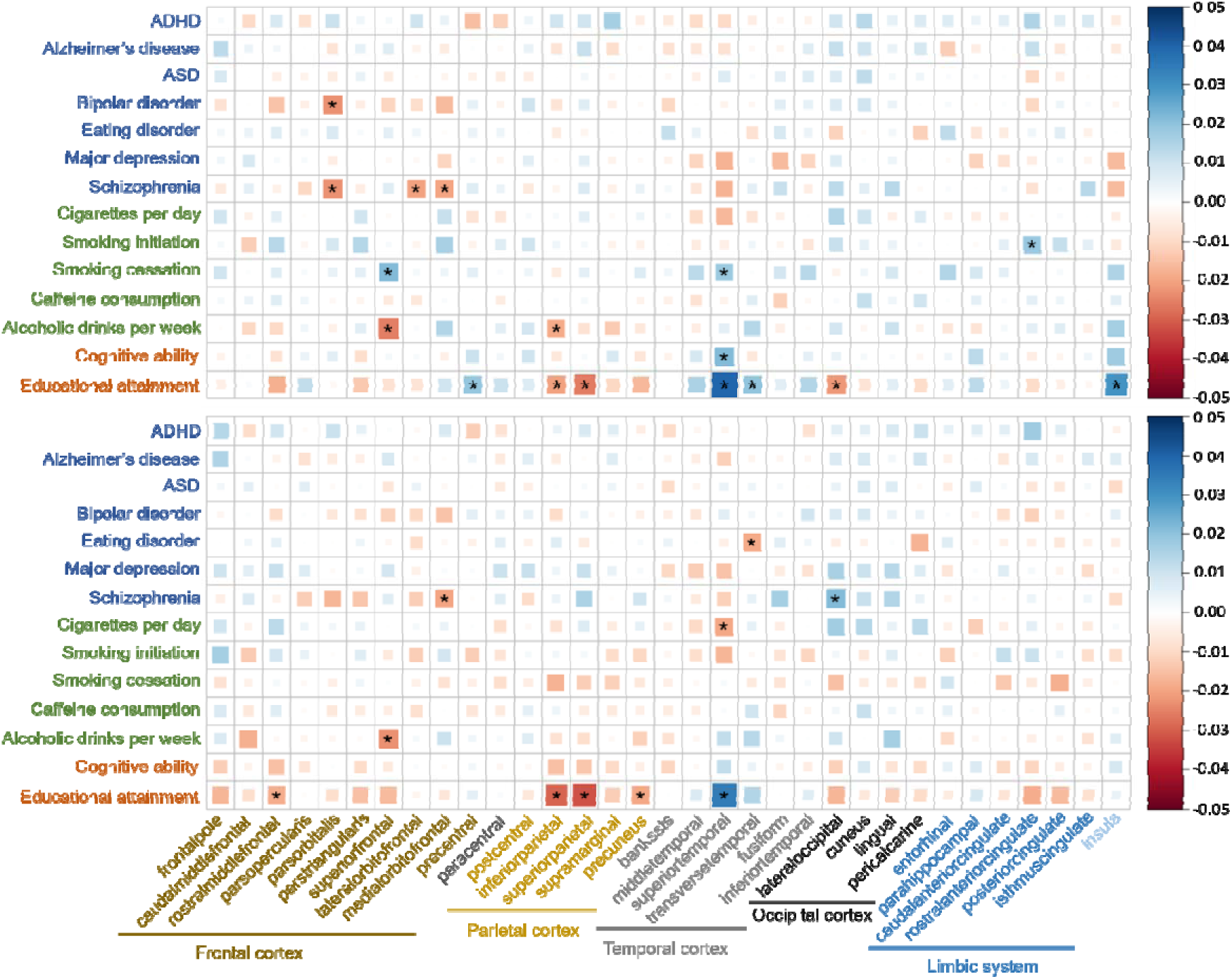
Associations between the 14 PGSs and the CT of 66 cortical regions, corrected for average CT. The size and color of each box indicate the magnitude of the association. Blue squares indicate positive associations, and red squares indicate negative associations. The upper half shows the PGS-CT correlation map across the left hemisphere, and the lower half across right hemisphere. Asterisks indicate significant associations after FDR multiple testing correction.

Associations without correction for global brain structure are presented in the supplementary materials (Supplementary Fig. 5-7, **Supplementary Tables 10-12**). Those results showed considerable overlap with those for global brain structure.

### Genetic relationships with brain function

UK Biobank used group independent component analysis (ICA) to decompose resting-state fMRI data into 21 major resting-state networks (Supplementary Fig. 8). Activity (temporal fluctuation) within these 21 networks and 210 FCs (temporal correlations) between these networks were then estimated for all subjects (see **Methods**). We examined the associations between the 14 PGSs and these 231 functional measures. Besides age, sex, MRI scanning positions, and genetic principal components, we accounted for the effects of mean resting-state fMRI head motion averaged across space and time points (**see Methods**).

We found 22 significant associations between PGSs and activity within 21 networks (Fig. 4, supplementary Fig. 9, **supplementary Table 13**). Of them, higher PGSs for bipolar disorder and schizophrenia were significantly associated with lower temporal fluctuations of the occipitotemporal cortex (node 2), cerebellum (node 15), basal ganglia (node 18), and posterior cingulate cortex (node 20) (Fig. 4D and 4G). The PGS for major depression showed significant negative associations with activity of the sensorimotor network (node 10, 11, 12) and cerebellum (node 15) (Fig. 4F). The PGS for educational attainment was negatively associated with activity of the visual cortex (node 2, 4), salience network (node 3), and frontal cortex/precuneus/para hippocampal gyrus (node 7) (Fig. 4N). In contrast, the PGS for smoking initiation showed a significant positive association with activity of the superior temporal gyrus (node 17) (Fig. 4I). Although the pattern of results was comparable, the genetic correlation analysis showed no significant associations between brain functional activity and the 14 mental health and cognitive traits (Supplementary Fig. 10, **Supplementary Table 14**).

**Fig 4.**
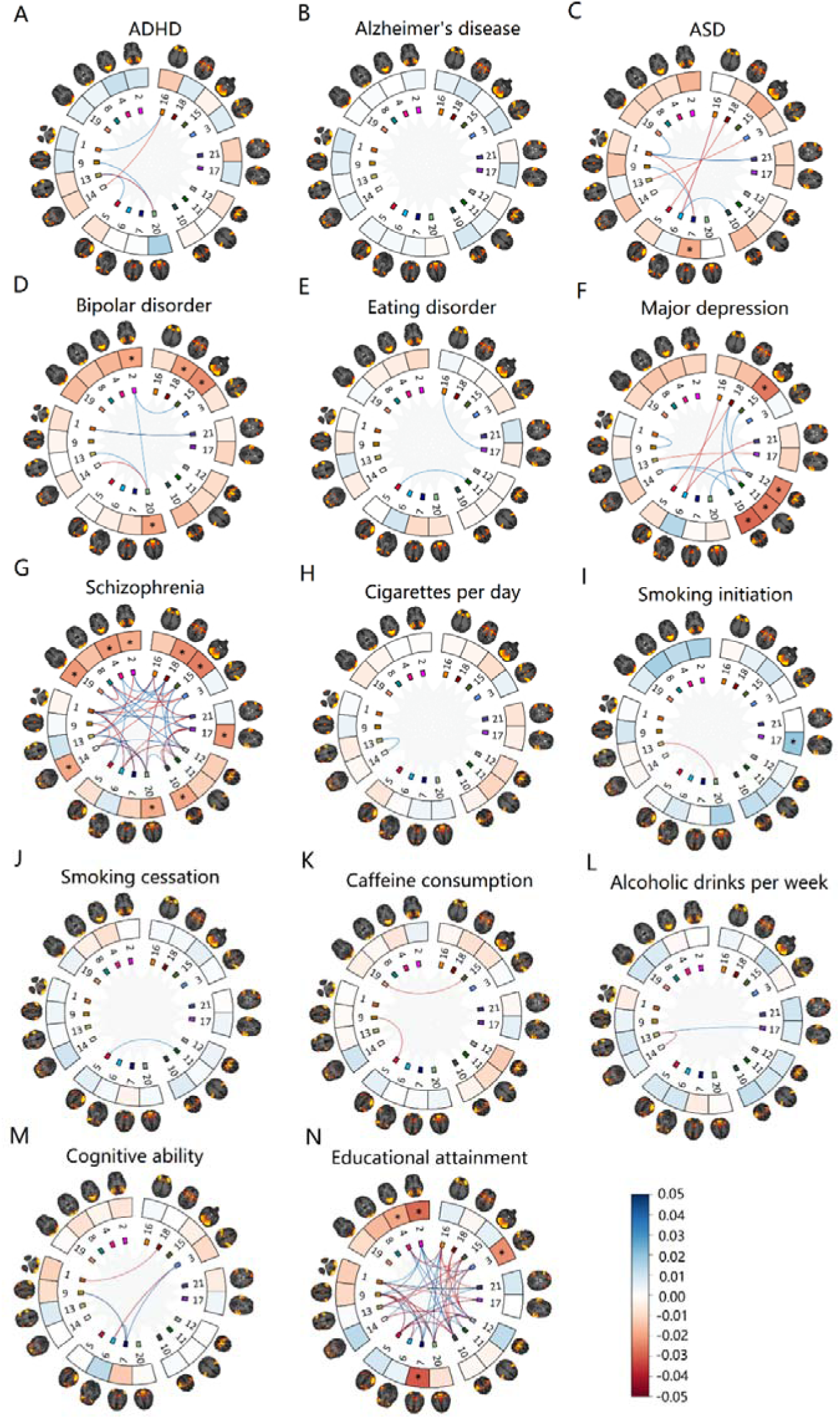
Significant associations between the 14 PGSs and functional measures. The nodes in each circle show the 21 resting-state networks. Asterisks indicate significant associations with activity (temporal fluctuation) of the 21 networks after FDR multiple testing correction. The lines between the nodes indicate significant FCs between the 21 functional networks after FDR multiple testing correction. The color of each line and node indicates the magnitude of the associations. Blue squares indicate positive associations, and red squares indicate negative associations.

A total of 133 significant associations between PGSs and FCs were observed (Fig. 4, **Supplementary Table 15**). PGSs for educational attainment and schizophrenia showed the largest number of significant associations with FCs (N = 46 and N = 43, respectively) (Fig. 4G and 4N). In addition, the PGS for cognitive ability showed 5 significant associations with FCs (Fig. 4M), and PGSs for various mental health disorders (ADHD, ASD, bipolar disorder, major depression) showed between 5 and 12 significant associations with FCs (Fig. 4A, C, D, F). The PGSs for Alzheimer’s disease, eating disorder, and the five substance use traits only showed 0-2 significant associations with FCs (Fig. 4B, E, H-L). The genetic correlation analysis only showed a few significant genetic correlations between FCs and schizophrenia, cognitive ability, and educational attainment (Supplementary Fig. 11, **Supplementary Table 16**).

### Multivariate modeling of joint relationships between PGSs and brain measures

The univariate analyses reported above showed that PGSs of multiple traits were associated with variability in both brain structure and function (e.g., educational attainment, cognitive ability, ADHD, major depression), whereas other PGSs were more specifically associated with either brain structure (e.g., Alzheimer’s disease, substance use traits) or brain function (e.g., other psychiatric disorders). In order to decompose the association patterns of various traits with brain structure and function, we performed a data-driven multivariate statistical analysis of the genetics and imaging data with split-half cross-validation on basis of partial least squares (PLS) regression^21^ (Fig. 5 **and Methods**). We focused on the first five components which ensured that we can get stable and robust latent variables^21^. This method with inherent replication identifies multivariate components that link multiple traits with multiple brain imaging variables.

**Fig 5.**
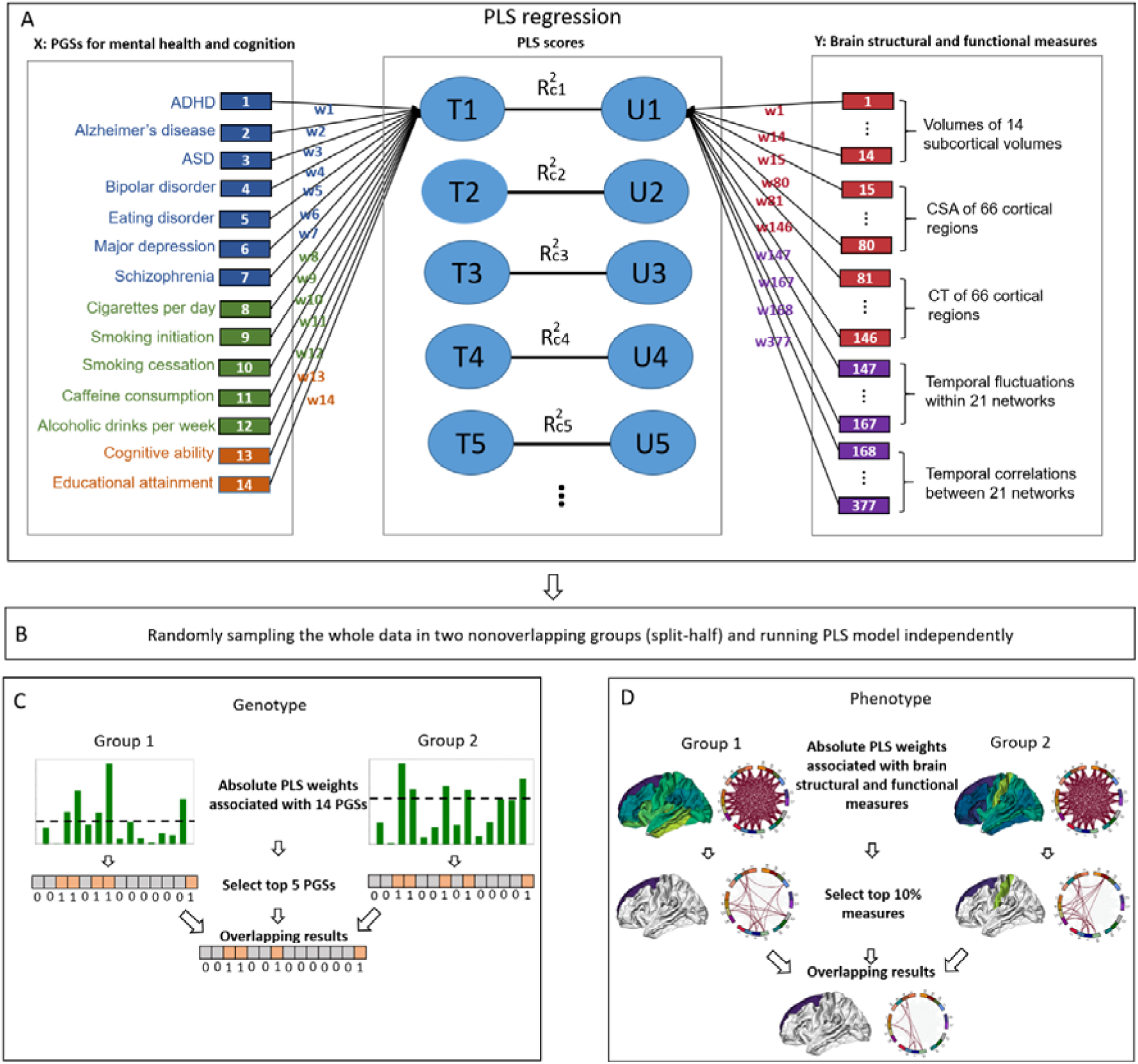
Flow chart of data-driven multivariate statistical analysis. (A) Input features of PLS regression included PGSs for 14 mental health and cognitive traits and 377 structural and functional measures (without three global structural brain measures). w_i_ indicated the weight of i-th input feature, which was served as the importance of relative importance for describing the multimodal relationship. Ti and Ui respectively indicated the i-th genotype and phenotype PLS component transformed from genetic and imaging data. (B) The dataset was randomly divided into two groups for validation in which PLS analyses were performed independently. This procedure was repeated 5000 times. The averaged results from the 5000 iterations identified the robust and consistently selected PGSs and brain imaging measures that received the largest weights in both groups for each iteration. (C) For each repetition, the absolute weights of 14 PGSs were sorted, and the top 5 PGSs were selected in two random groups. The selected PGSs were labeled one or zero otherwise. The resulting binary arrays independently estimated in the two groups were then merged, such that only replicated PGSs receiving the largest PLS weights were selected. (D) For each repetition, the absolute weights of brain measures were sorted, and the top 10% brain measures are selected in two random groups. The selected brain measures are labeled one or zero otherwise. The resulting binary arrays independently estimated in the two groups were then merged, such that only replicated brain measures receiving the largest weights were selected.

Fig. 6A and 6B respectively show the mean weights for the first PLS genotype and phenotype component, which reflect the importance of each input variable for the PGSs-imaging relationships in the PLS model. Fig. 6C and 6D show the probabilities for PGSs and imaging measures receiving the largest PLS weights in two random groups across 5000 repetitions. PGSs for ADHD, smoking initiation, cognitive ability, and educational attainment had higher weights and were consistently selected for the first genotype component. The corresponding imaging component was mainly associated with larger CSA of many cortical regions, especially the frontal and temporal regions, as well as larger volume of the thalamus, whereas regional CT and functional measures had low weights and were not consistently selected. Similarly, Fig. 6E-H show the weights and probabilities for the second PLS genotype and phenotype component. The genetic patterns of PGSs for psychiatric disorders (ASD, bipolar disorder, major depression, and schizophrenia) corresponded consistently to lower activity of 5 functional networks and to 10 both stronger and weaker FCs. All structural measures had low weights and were not consistently selected. The selected nodes (resting-state networks) included the visual system (node 2 and 4), somatosensory system (node 10), cerebellum (node 15), and basal ganglia (node 18). FCs included 5 positive and 5 negative associations across the entire brain, including networks often associated with psychiatric disorders such as the default mode network, salience network, and ventral and dorsal attention networks. The PGSs and brain measures were not stably and consistently selected across 5000 repetitions in the last three components, so the weights and probabilities of these measures were shown in the supplementary materials (Supplementary Fig. 12-13, **Supplementary Table 17**).

**Fig 6.**
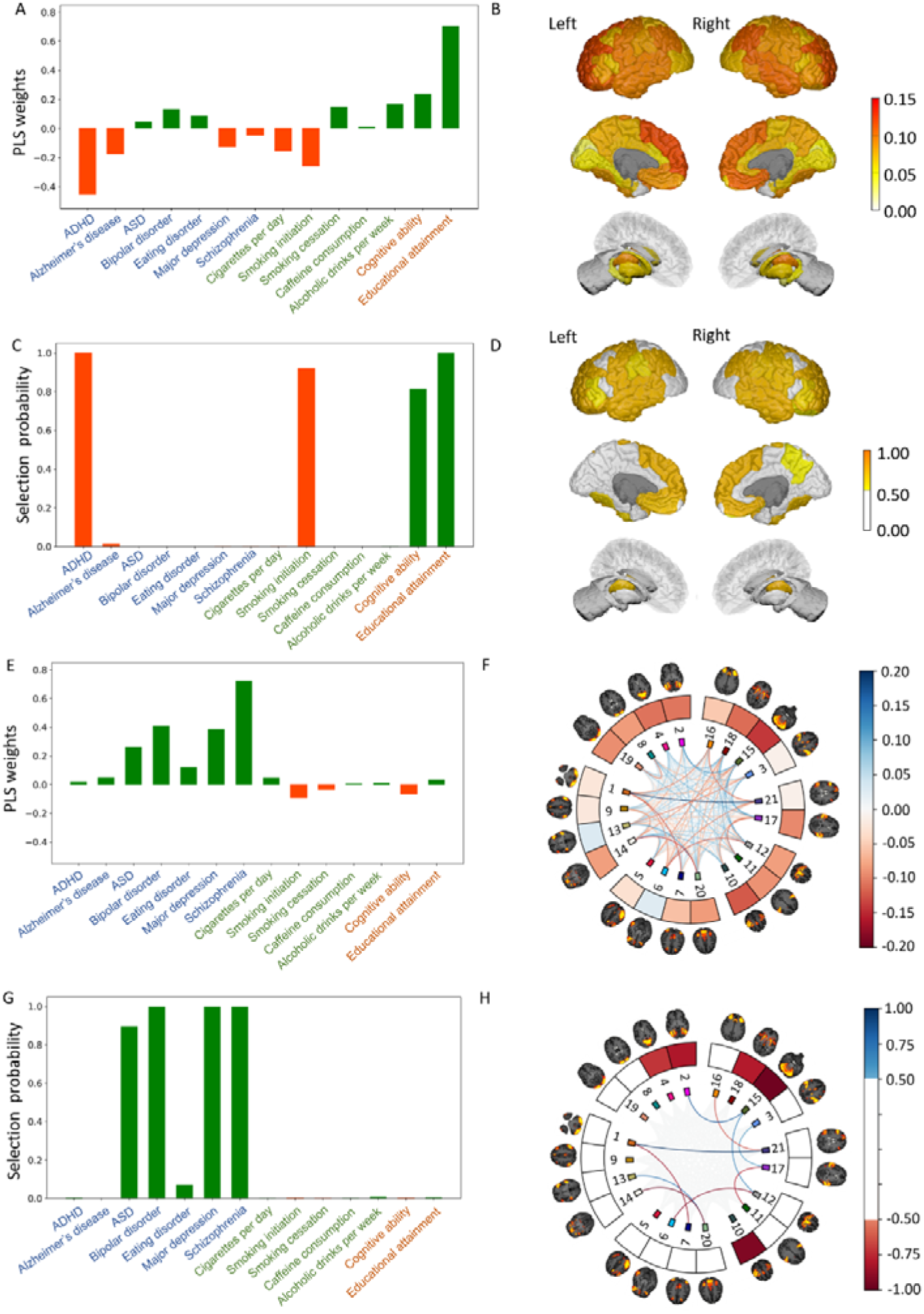
First two components of the PLS model. (A) The mean weights across 5000 repetitions for 14 PGSs in the first genotype component. (B) The mean weights across 5000 repetitions for the CSA of cortical regions and volumes of subcortical regions in the first phenotype component. (C) Probabilities of the 14 PGSs of being selected across 5000 repetitions in the first genotype component. (D) Probabilities of regional CSA and subcortical volumes of being selected across 5000 repetitions in the first phenotype component. (E) The mean weights across 5000 repetitions for 14 PGSs in the second genotype component. (F) The mean weights across 5000 repetitions for activity of 21 functional networks and 210 FCs between these networks in the second phenotype component. (G) Probabilities of the 14 PGSs of being selected across 5000 repetitions in the second genotype component. (H) Probabilities of activity of 21 functional networks and 210 FCs between these networks of being selected across 5000 repetitions in the second phenotype component.

## Discussion

We systematically investigated the genetic relationships of the 14 mental health and cognitive traits with 380 structural and functional brain measures using neuroimaging and genetic data from 36,799 participants. The results show that genetic predispositions to ADHD, smoking initiation, cognitive ability and educational attainment are more strongly associated with brain structure than with brain function, whereas genetic predispositions to most psychiatric disorders (ASD, bipolar disorder, major depression, and schizophrenia) are more strongly associated with brain function than with brain structure. To our knowledge, this is the first study providing evidence for these differential associations, and suggests that there are distinct neural pathways through which genes eventually influence mental health and cognition.

Mass univariate correlation analyses showed various and complex associations between the14 PGSs and 380 brain measures. For global structural measures, PGSs for ADHD, Alzheimer’s disease, major depression, eating disorder, smoking traits, alcohol consumption, and cognitive traits were significantly associated with total CSA or ICV, whereas PGSs for several psychiatric disorders (ASD, bipolar disorder, and schizophrenia) showed no significant association. For regional structural measures, only the PGS for educational attainment showed widespread significant associations. For functional measures, PGSs for cognitive traits and psychiatric disorders showed widespread significant associations with activity of functional networks and FCs between networks.

We also estimated the SNP-based genetic correlations between these traits and neuroimaging features and found generally consistent results, although fewer associations were significant due to lower power of these analyses. A recent study compared the genetic correlations of different traits with brain structural measures and also reported that global cortical structure is genetically correlated with cognitive function, depression, and ADHD^8^. This study however did not account for the effects of global measures in its regional analyses, which affects its evidence for the specific roles of cortical regions in genetic mechanisms of various traits.

In order to decompose the joint genetic association patterns into underlying components, we performed a data-driven multivariate statistical analysis (PLS regression) of the genetics and imaging data. The first PLS component confirmed that ADHD was more strongly associated with brain structure than with brain function. In fact, previous studies consistently reported significant genetic sharing between ADHD and total CSA and ICV^8, 23, 24^, whereas only a few FCs were significantly associated with the PGS for ADHD^25^. Epidemiologic and clinical studies have found that genetic risk factors that affect the structure and functional capacity of brain networks are involved in the etiology of ADHD^26, 27^. Many genes that have been associated with ADHD, such as FOXP2, SORCS3, and DUSP6, are highly expressed in the brain and implicated in neurodevelopmental processes^26^.

Comparable results were obtained for smoking initiation, which showed a similar association pattern with the brain measures as ADHD. Although the molecular mechanisms underlying these associations have yet to be elucidated, a previous gene set analysis of enriched biological pathways provided some possible clues that brain dopamine receptor signaling is associated with smoking initiation^28^. For example, BDNF regulates synaptic plasticity and survival of cholinergic and dopaminergic neurons^29^. Genetic variants within the BDNF gene were found to be genome-wide significantly associated with smoking initiation^30^ and the gene was highly expressed in the prefrontal cortex, which might be implicated in the cognitive-enhancing effects of nicotine^31^. This is in line with our finding that the PGS for smoking initiation was especially associated with the CSA of several frontal regions.

In addition, genetic predispositions to cognitive traits were significantly associated with both brain structure and function, though the first PLS component showed that the association with brain structure was stronger. Our results for cognitive traits are in line with previous studies in which the PGS for educational attainment was associated with larger brains^10, 32^ and CSA and CT in some frontal and temporal regions mediated the association between the PGS for intelligence and the *g*-factor^11^, but no large-scale study provided credible and sufficient evidence on relationships of PGSs for cognitive traits with brain function. Previous studies have presented some possible explanations for the associations, describing that biological annotation of genes associated with educational attainment were involved in brain-development processes and neuron-neuron communication^33, 34^.

The second PLS component shows that PGSs for most psychiatric disorders (ASD, bipolar disorder, major depression, and schizophrenia) were more strongly associated with brain function than with brain structure. Previous studies for the genetic relationships between psychiatric disorders and global brain structure produced mixed results, likely because of limited sample sizes. For example, some studies reported that a decrease in total brain volume or cortical thickness was significantly associated with an increased polygenic risk for schizophrenia^17, 35^, whereas another study with a relatively large sample found no significant associations^16^. With respect to brain function, previous studies indicated that PGSs for ASD, bipolar disorder, major depression, and schizophrenia were associated with functional measures^5, 36, 37^. Overall, although our results do support a limited link between PGSs for psychiatric disorders and brain structure, results from our multivariate analysis highlights that the association with brain function is stronger when both are considered together. The stronger associations between PGSs and brain function may indicate that genetic predispositions to psychiatric disorders are likely to cause difficulties in information integration across the brain, which in turn makes individuals more prone to developing a psychiatric disorder^13^.

We note several limitations to our study. First, accuracy of the PGSs highly depends on the sample size of the discovery GWAS summary statistics. Thus, the statistical power differs substantially between different traits, which may influence the comparisons of association patters. Second, our results point to genetic associations between different traits and brain measures, but the nature of the association is not yet known. We cannot distinguish genetic pleiotropy versus casual relationships from mental health and cognitive traits to brain structure and function or vice versa. Thirdly, the polygenic scores may contain some environmental effects, because genetic effects may possibly act via the environment to influence the brain. These environmental effects can include the influence of parents who provide the offspring with their genome as well as with their rearing environment^38^, which can result in an inflation of estimated genetic effects. Gene-environment correlations can also occur on a regional level: the educational attainment polygenic score, for example, has been shown to influence where people live (individuals with a higher polygenic score are more likely to migrate to a richer neighborhood), which may have an effect on mental health and cognitive outcomes^39^.

In summary, this study provides evidence for distinct association patterns of genetic predispositions to mental health and cognition with brain structure and function, with genetic predispositions to ADHD, smoking initiation, and cognitive traits having stronger associations with brain structure than with brain function, whereas genetic predispositions to most other psychiatric disorders having stronger associations with brain function than with brain structure. These results bring us closer to understanding the neurobiological basis of these traits.

## Data Availability

Data is available from UK Biobank

https://www.ukbiobank.ac.uk/

## ACKNOWLEDGMENTS

This research has been conducted using data from UK Biobank under application numbers 40310 and 30091. S.L.is supported by the China Scholarship Council. A.A. & K.J.H.V. are supported by the Foundation Volksbond Rotterdam.

## METHODS

### Participants

The data used in this study are from UK Biobank^22^. We used the latest available imaging release (on January 2020) including 36,799 European ancestry participants from across the United Kingdom with both genetic and imaging data (19,512 females and 17,287 males, mean age = 54.90 years, SD = 7.43 years, range from 40 to 70 years). The National Health Service North West Centre for Research Ethics Committee provided UK Biobank project with ethical approval (reference: 11/NW/0382) (http://biobank.ctsu.ox.ac.uk/crystal/field.cgi?id=200).

### Imaging procedures

The UK Biobank study contained 6 different kinds of brain imaging measures (http://biobank.ndph.ox.ac.uk/showcase/label.cgi?id=100)^40^. For this study, we used the T1 structural and resting-state functional MRI (rsfMRI) measures to represent brain structure and function, respectively. For brain structure, there were three global measures (total cortical surface area (CSA), intra-cranial volume (ICV) and average cortical thickness (CT)). Apart from them, we also focused on the regional measures including the volumes of 14 subcortical regions, 66 surface area of parcels identified on the cortical surface, and cortical thickness within these areas. The cortical parcellations were based on the Desikan-Killiany (DK) atlas^41^. For brain function, there were activity within 21 functional networks and functional connectivities (FCs) between these networks derived from rsfMRI. Briefly, group independent component analysis (group-ICA) was applied to decompose the rsfMRI data into a specified number of networks based on more than 4000 participants by UK Biobank^42^ (https://www.fmrib.ox.ac.uk/ukbiobank/). Then, the averaged time series of different brain regions (ICA components) were extracted to estimate the fluctuation of amplitudes (temporal standard deviation) as the measures for functional activity. In addition, the temporal correlations of averaged time series between different pairs of regions ware estimated as the FC. This study used the partial correlation matrix on 25 dimensions provided by UK Biobank. Detailed information about the image acquisition protocols and processing pipeline is provided at: http://biobank.ctsu.ox.ac.uk/crystal/crystal/docs/brain_mri.pdf.

### Genotype data

Participants provided their blood samples when they visited a UK Biobank assessment center, From the blood sample, DNA was extracted^43^ and genotyping was carried out by Affymetrix Research Services Laboratory using the Applied Biosystems UK BiLEVE Axiom Array or Applied Biosystems UK Biobank Axiom Array. These two arrays were very similar sharing 95% of marker content, and only the markers included in both arrays were considered for later genotype processing. Quality control (QC) procedures are described elsewhere^44^. In short, 1,252,123 SNPs remained after filtering out with MAF < .01 and missingness > .05. We then filtered out individuals with non-European ancestry and repeated the SNP QC on unrelated Europeans, leaving 1,246,531 SNPs. Furthermore, to attenuate population structure confounds to genetic analysis, we excluded individuals with non-European ancestry, and performed a principal component analysis (PCA) to capture ancestry differences within the white British population^44^. We kept the top 25 principal components (PCs) as covariates for later analysis.

### Polygenic scores (PGSs)

PGSs were calculated by summing the number of risk alleles an individual carries weighted by the strength of the association of each allele with a specific trait. The strength of association for each SNP is based on the effect size estimate from existing GWAS summary statistics (**Supplementary Table 1**). To avoid overestimation of the genetic predisposition estimate, the discovery samples used for GWAS of a trait should not include the target sample (here UK Biobank dataset). In this study, we created PGSs for 14 mental health and cognitive traits. In detail, PGSs were constructed using the summary-data based best linear unbiased prediction (SBLUP) approach^45^. This method maximizes the predictive power by creating scores with best linear unbiased predictor properties that account for linkage disequilibrium (LD) between SNPs^39^. As a reference sample for the LD, we used the random sample of 10,000 unrelated individuals from UK Biobank that were imputed using the Haplotype Reference Consortium (HRC) reference panel^46^. The polygenic scores were computed on a set of 1,312,100 autosomal HapMap 3 SNPs with a minor allele count of >5, an info score of >0.3, Hardy–Weinberg equilibrium (HWE) P-value (P_HWE_) < 10^−6^ and missingness < 0.05.

### Associations between the PGSs and brain measures

We firstly investigated the associations between PGSs for the 14 traits and global brain measures (total CSA, ICV, and average CT). We included age, sex, age*sex, age^2^, age^2^*sex, head positions (X, Y, Z), and the top 25 genetic PCs as covariates^47^. The 14 PGSs and 3 global brain measures were then regressed out of these covariates. Next, we calculated the Pearson correlation coefficients among regressed values (14 PGSs x 3 global measures) using the SciPy Stats package in python.

PGSs for the 14 traits were then tested for associations with the volumes of 14 subcortical regions as well as CSA and CT for the 66 DK atlas regions. For these analyses, ICV (for subcortical volumes), total CSA (for regional CSA), or average CT (for regional CT) were added as covariates to account for the global brain structural effects. When regressed out of the covariates respectively, Pearson correlation coefficients among regressed values (14 PGSs x 146 regional structural measures) were calculated using the same method.

Finally, we investigated the associations between PGSs for the 14 traits and brain functional measures which includes activity of 21 functional networks and 210 FCs between these networks. Besides age, sex, head positions and genetic principal components, we accounted for the effects of the head motion in the analysis. The same statistical procedures were conducted based on 14 PGSs x 231 functional measures.

We applied false discovery rate (FDR) multiple testing correction with a significance threshold of 0.0167 (0.05/3) to account for the above three sets of analyses: global structural measures, regional structural measures, or functional measures.

### Genetic correlation analysis

LD score regression^20^ was used to calculate the genetic correlations of the brain imaging measures with mental health disorders, substance use, or cognitive traits. First, we used mixed linear model (MLM)-based approaches to conduct the GWASs for 380 brain measures in UK Biobank using fastGWA^48^. To control for relatedness, we included a genetic relatedness matrix (GRM) in the model. In addition, covariates like those applied in the Pearson correlation analysis were regressed out.

The LD-SCORE regression software (v1.0.0; https://github.com/bulik/ldsc) was then used to estimate SNP-based heritability for the brain measures and to compute the genetic correlations of these brain measures with the 14 mental health and cognitive traits, using GWAS summary statistics^49^. In short, the genetic correlation between traits is based on the estimated slope from the regression of the product of z scores from two GWASs on the linkage disequilibrium score, and represents the genetic covariation between two traits based on all of the polygenic effects captured by the included SNPs^39^. The genome-wide linkage disequilibrium measurements used by these methods were taken from the 1000 Genomes Project^50^ (European populations from the HapMap 3 reference panel).

### Joint partial least squares modeling analysis

To test for differences in relationships of 14 mental health and cognitive traits with brain structure and function, we applied joint partial least squares (PLS) modeling of the genetic and imaging modalities, which find the multidimensional direction in the genetic space that explains the maximum multidimensional variance direction in the imaging space^51^. We performed a canonical symmetric version of the PLS regression from the scikit-learn package in python^52^ to find linear relations between two multivariate datasets (Fig. 5A). PLS is a method that resembles canonical correlation analysis^53^. Both of them are widely used to extract features from two sets of multi-dimensional variables. The fundamental difference between them is that the former estimates the latent components that maximize the global covariance while the latter maximizes the correlation. The detailed procedures of PLS method are represented in supplementary Fig 14^54^. In short, the input features included PGSs for the 14 mental health and cognitive traits as one set of variables and 377 structural and functional measures as the other. PLS modeling produces a weight for each input feature that represents its relative importance for describing the global joint multimodal relationship^21^. Comparing the weights helps identify PGSs that are linked to the patterns of brain structure or function. Age, sex, age*sex, age^2^, age^2^*sex, head positions, and the top 25 genetic PCs were added as covariates.

A robust approach for the stable estimation and interpretation of PLS weights was applied in this study (Fig. 5B-D), following a previously described procedure^21^. Briefly, a stability selection procedure was conducted to assess the reproducibility and robustness of the PLS parameters. The whole sample was randomly split in half and the PLS model was run on each sampled subgroup independently. Through 5000 repetitions of this procedure, we estimated a confidence measure for each input feature ranging between 0.0 and 1.0, indicating its probability containing highly reproducible PLS weights. Therefore, the measure can be regarded as the importance of input feature. Similar to the study by Lorenzi and Altmann^21^, the number of components was also set as five.

---

**Supplementary Fig 1:**
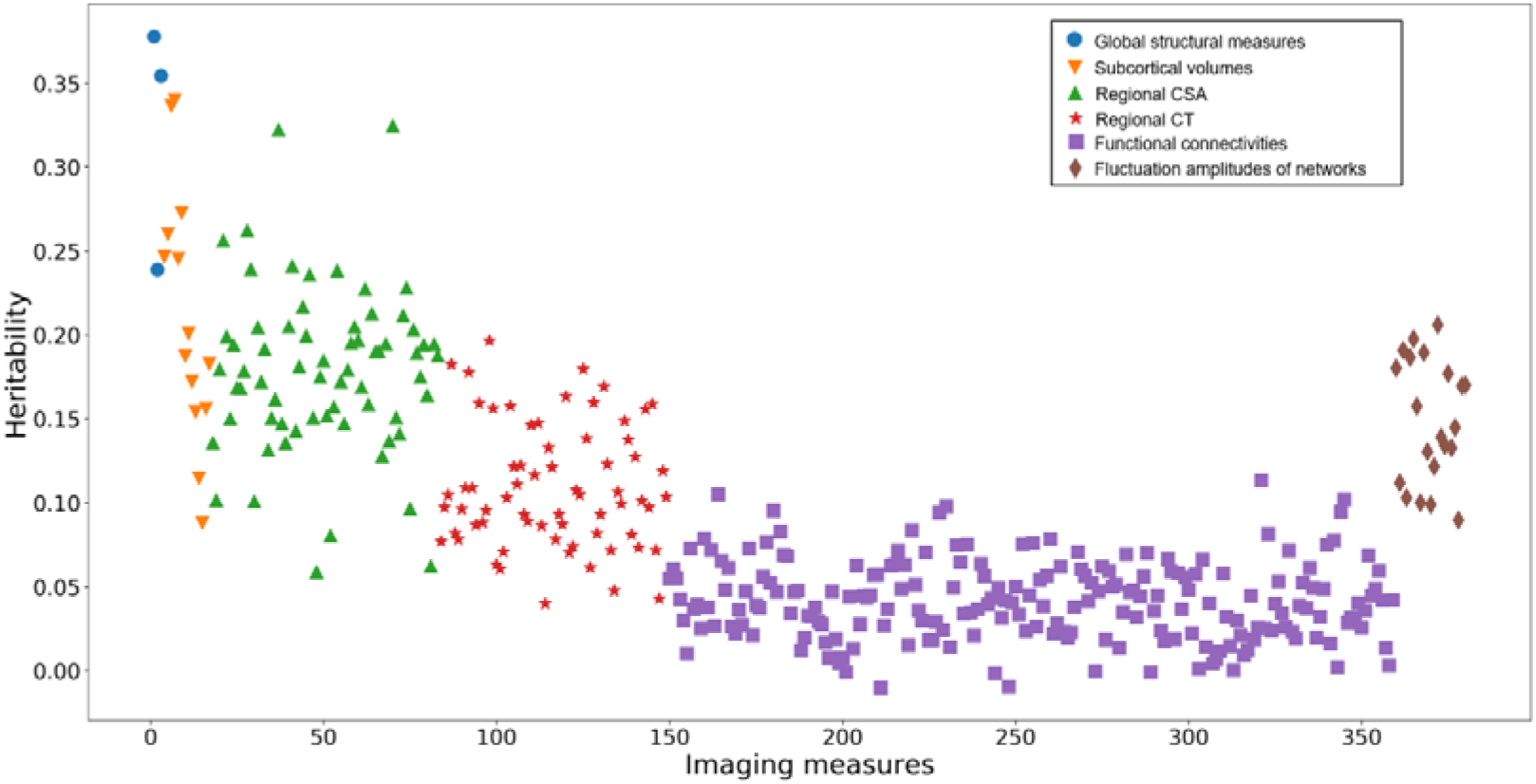
Estimated SNP Heritability of 380 structural and functional neuroimaging measures.

**Supplementary Fig 2:**
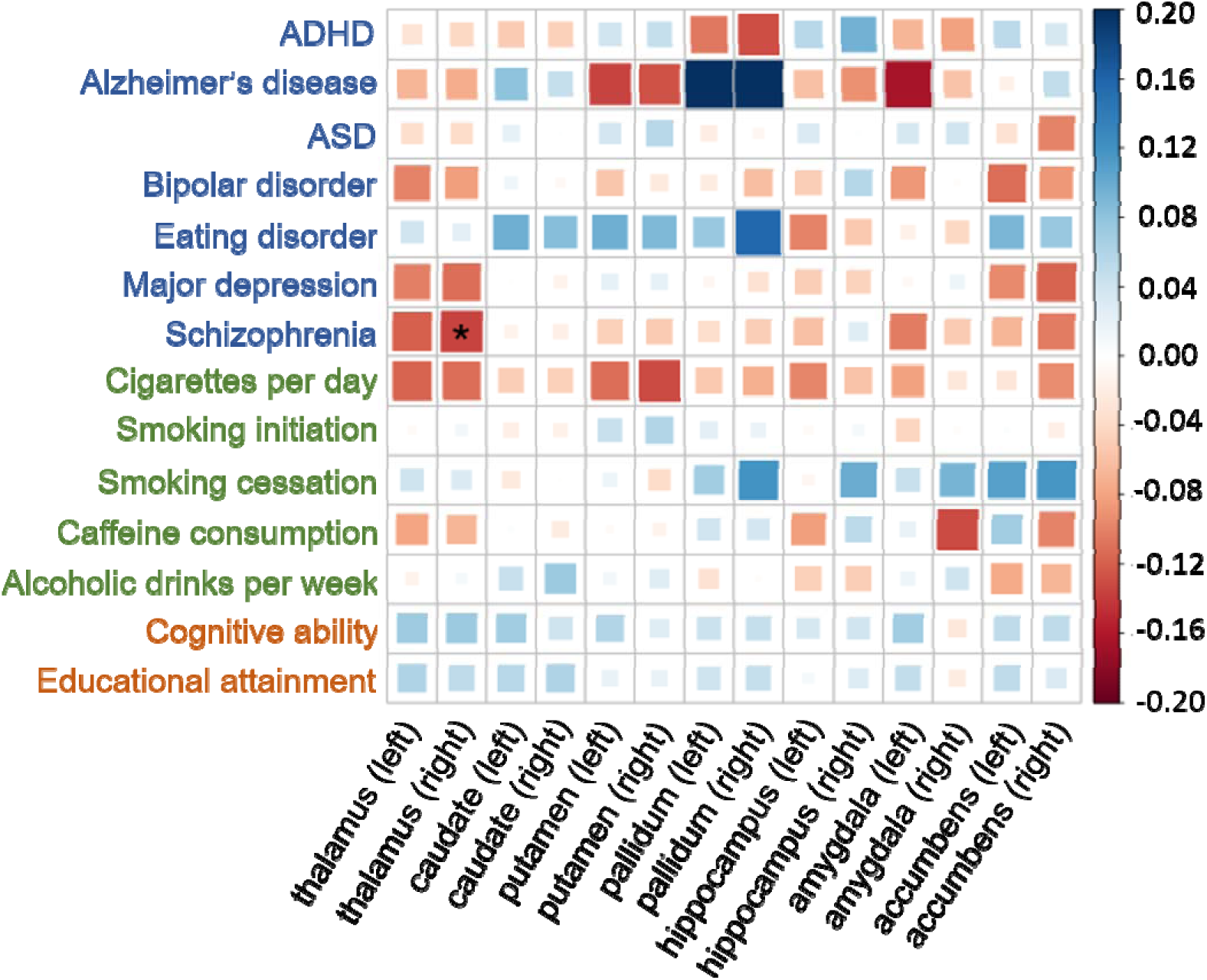
Genetic correlations between the 14 PGSs and the 14 subcortical volumes, corrected for ICV. The size and color of each box indicate the magnitude of the correlation. Blue squares indicate positive correlations, and red squares indicate negative correlations. Asterisks indicate significant correlations after FDR multiple testing correction.

**Supplementary Fig 3:**
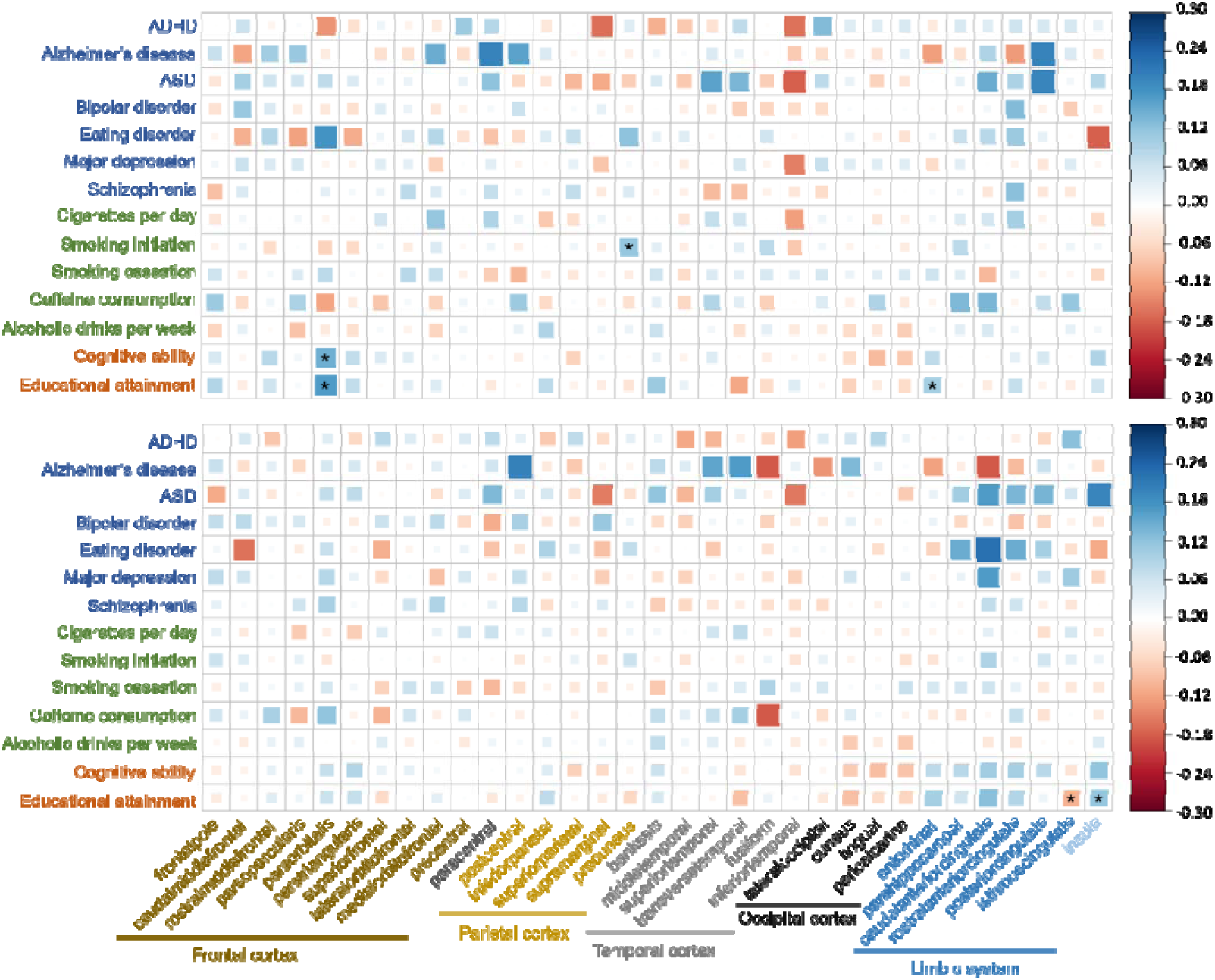
Genetic correlations between the 14 PGSs and the CSA of 66 cortical regions, corrected for total CSA. The size and color of each box indicate the magnitude of the correlation. Blue squares indicate positive correlations, and red squares indicate negative correlations. The upper half shows the correlation map across the left hemisphere, and the lower half across right hemisphere. Asterisks indicate significant correlations after FDR multiple testing correction.

**Supplementary Fig 4:**
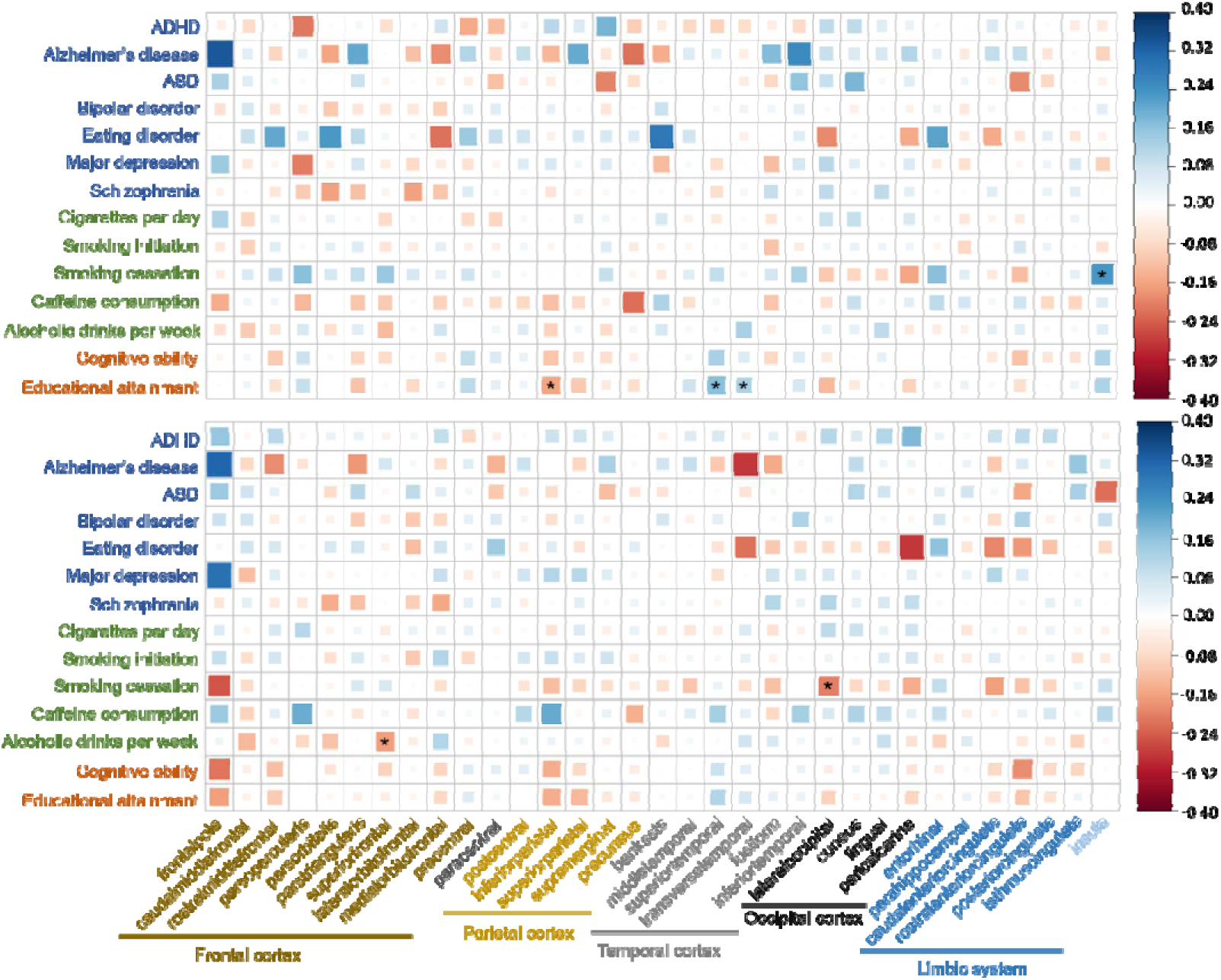
Genetic correlations between the 14 PGSs and the CT of 66 cortical regions, corrected for average CT. The size and color of each box indicate the magnitude of the correlation. Blue squares indicate positive correlations, and red squares indicate negative correlations. The upper half shows the correlation map across the left hemisphere, and the lower half across right hemisphere. Asterisks indicate significant correlations after FDR multiple testing correction.

**Supplementary Fig 5:**
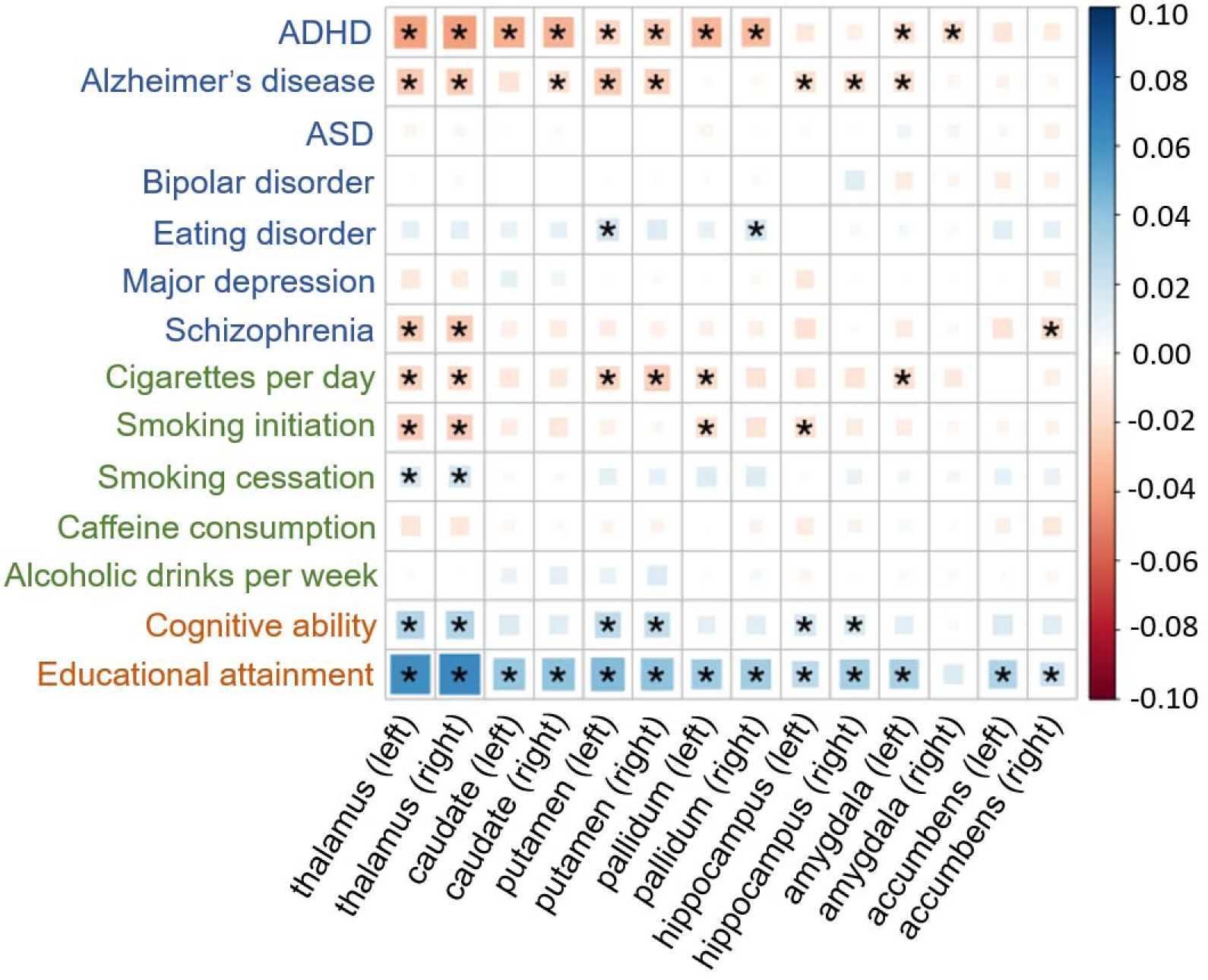
Associations between the 14 PGSs and 14 subcortical volumes, **not** corrected for ICV. The size and color of each box indicate the magnitude of the association. Blue squares indicate positive associations, and red squares indicate negative associations. Asterisks indicate significant associations after FDR multiple testing correction. Blue traits = mental health, green = drug use, red = cognition.

**Supplementary Fig 6.**
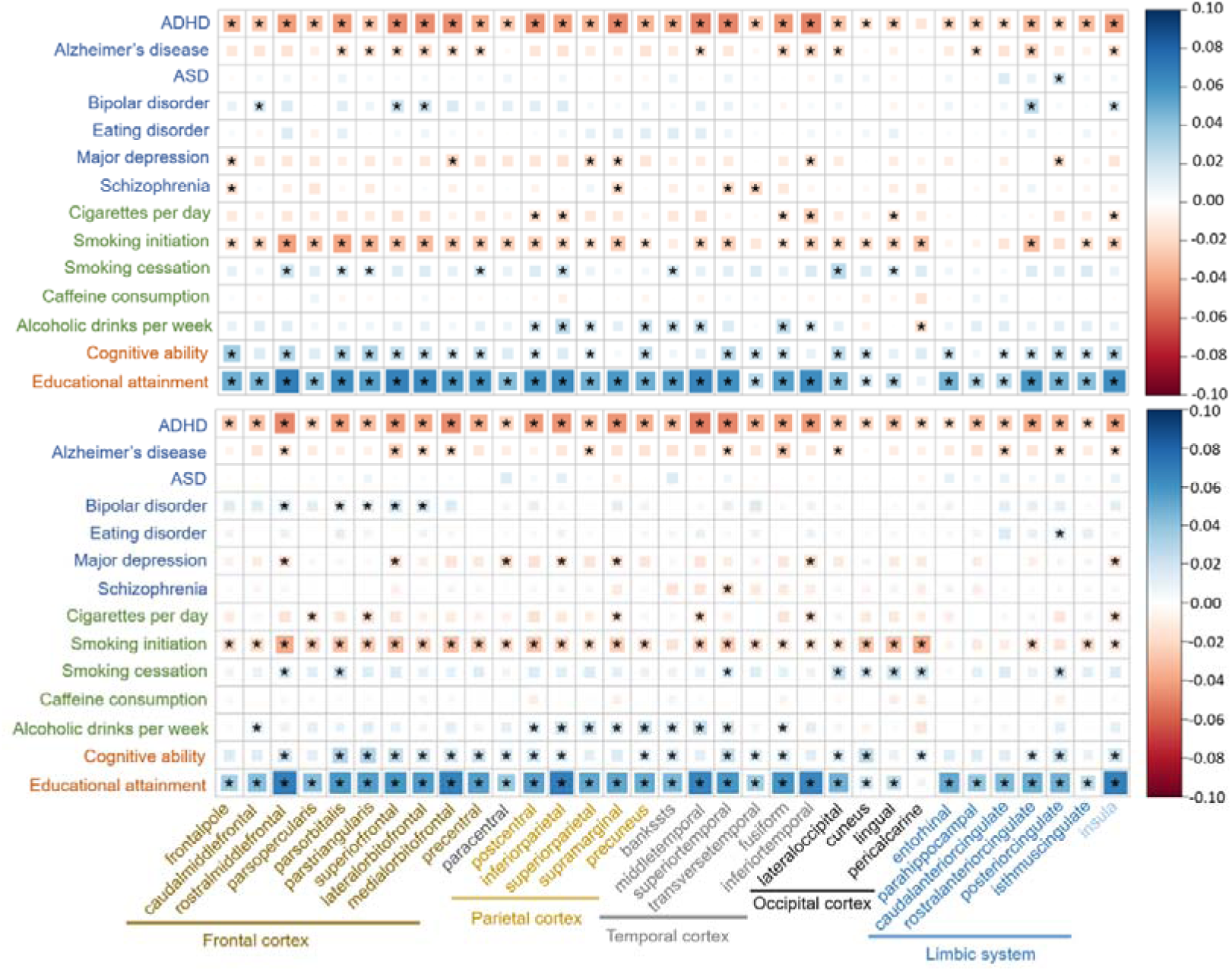
Associations between the 14 PGSs and the CSA of 66 cortical regions, **not** corrected for total CSA. The size and color of each box indicate the magnitude of the association. Blue squares indicate positive associations, and red squares indicate negative associations. The upper half shows the PGS-CSA correlation map across the left hemisphere, and the lower half across right hemisphere. Asterisks indicate significant associations after FDR multiple testing correction.

**Supplementary Fig 7.**
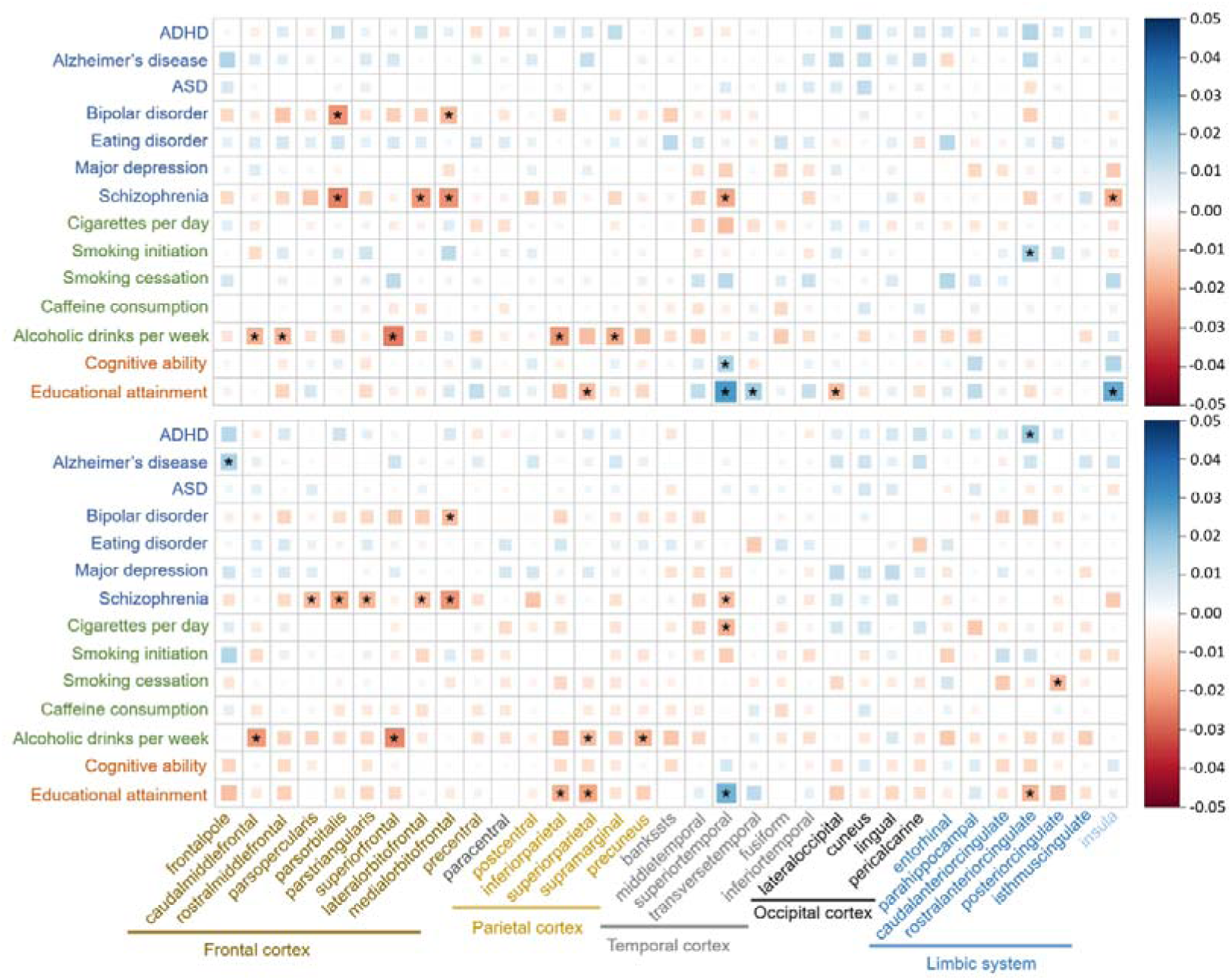
Associations between the 14 PGSs and the CT of 66 cortical regions, **not** corrected for average CT. The size and color of each box indicate the magnitude of the association. Blue squares indicate positive associations, and red squares indicate negative associations. The upper half shows the PGS-CT correlation map across the left hemisphere, and the lower half across right hemisphere. Asterisks indicate significant associations after FDR multiple testing correction.

**Supplementary Fig 8:**
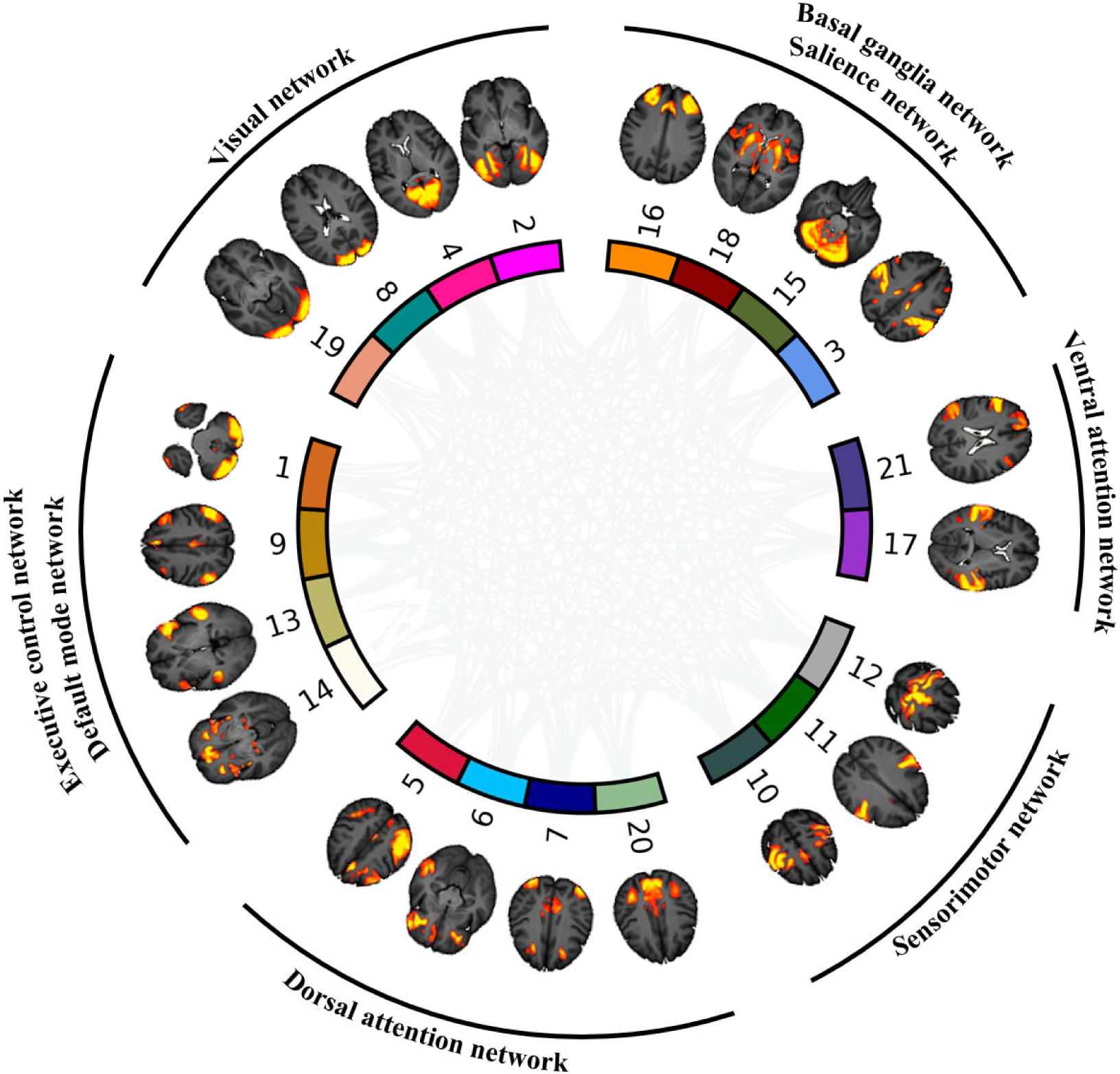
21 resting-state functional networks from group ICA analysis by UK biobank based on more than 4000 UK Biobank participants.

**Supplementary Fig 9:**
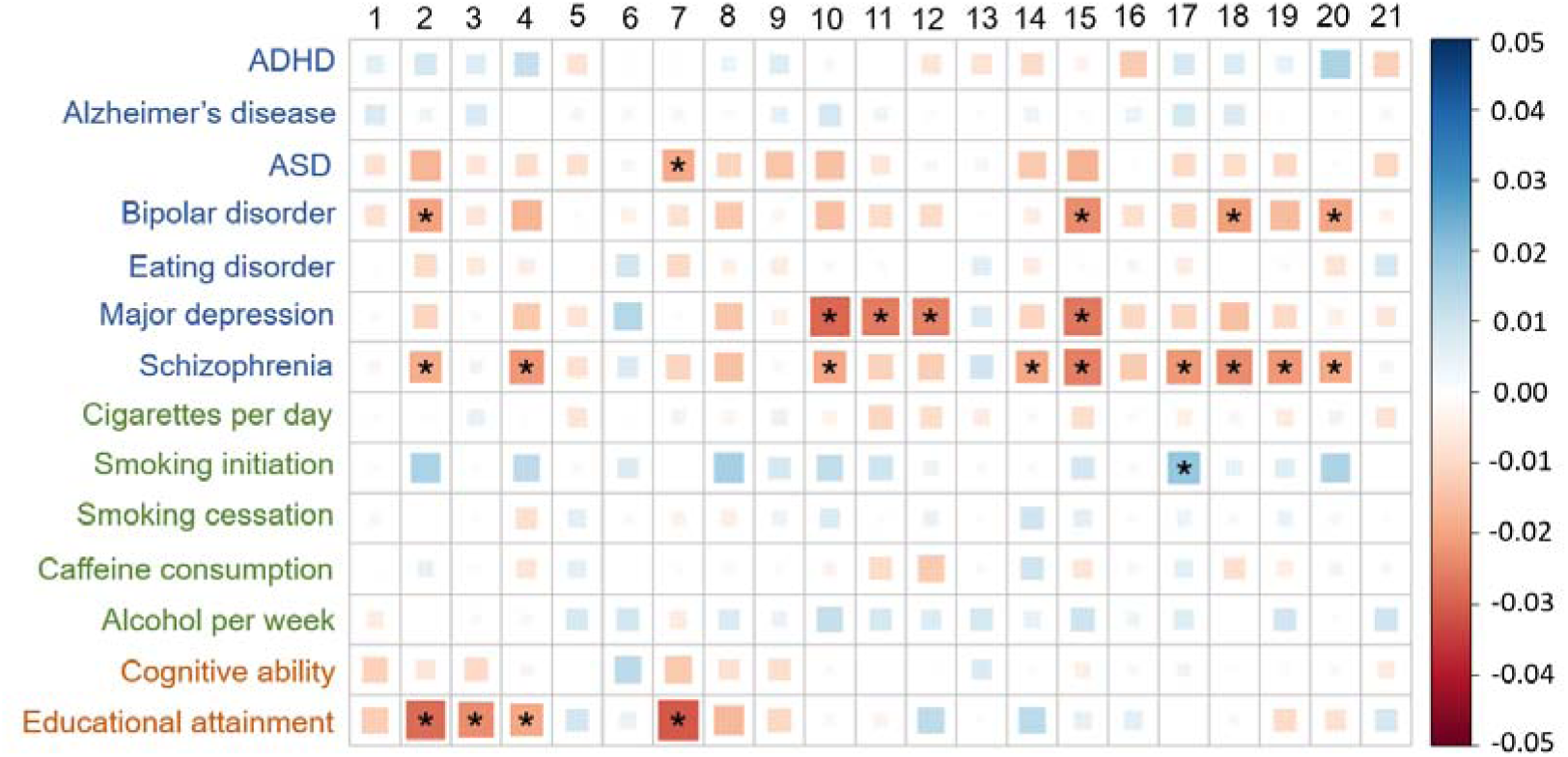
Associations between the 14 PGSs and activity (temporal fluctuation) of 21 functional networks. The size and color of each box indicate the magnitude of the association. Blue squares indicate positive associations, and red squares indicate negative associations.

**Supplementary Fig 10:**
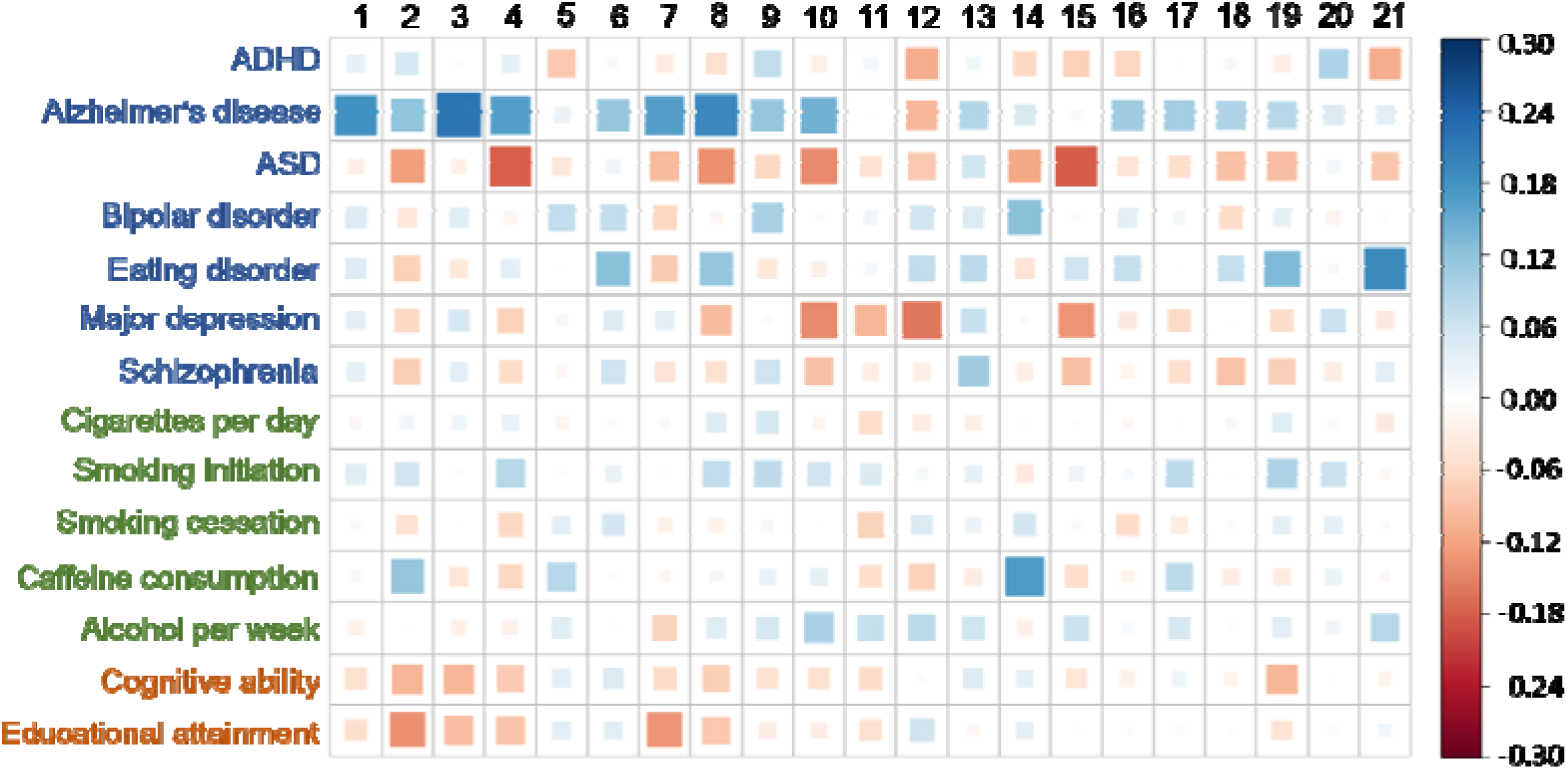
Genetic correlations between the 14 PGSs and activity (temporal fluctuation) of 21 functional networks. The size and color of each box indicate the magnitude of the correlation. Blue squares indicate positive correlations, and red squares indicate negative correlations.

**Supplementary Fig 11:**
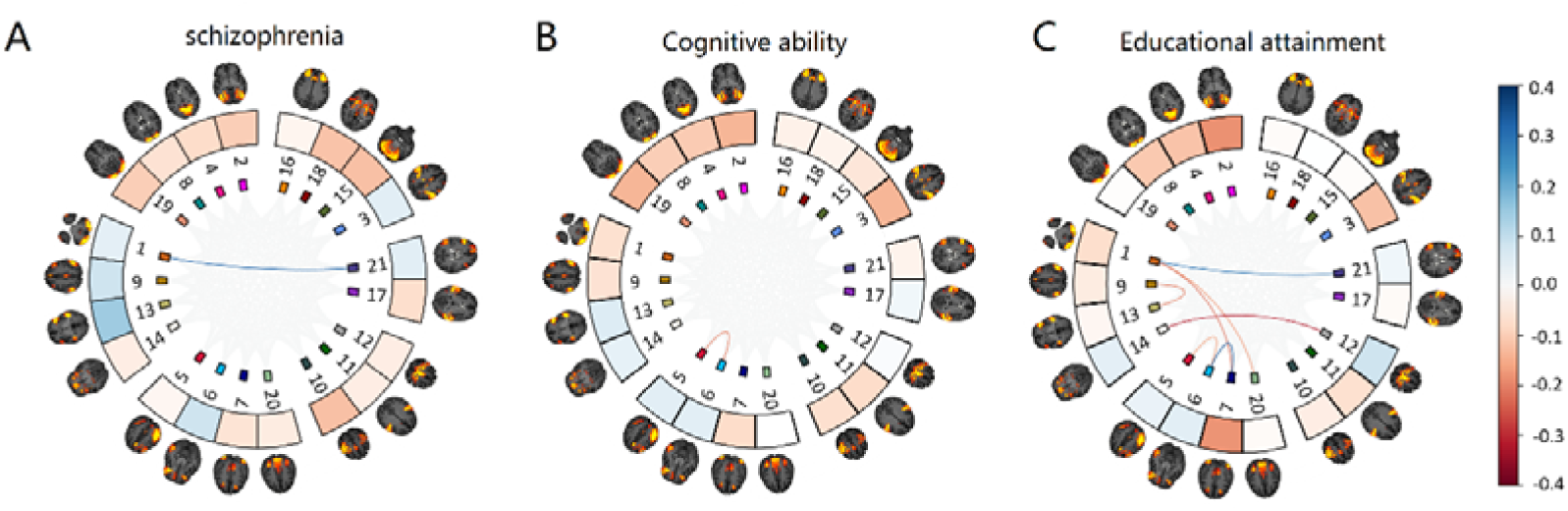
Significant genetic correlations between the 14 PGSs and FCs of 21 functional networks. The lines between the nodes indicate significant FCs between the 21 functional networks after FDR multiple testing correction. The color of each line indicates the magnitude of the correlations. Blue squares indicate positive correlations, and red squares indicate negative correlations.

**Supplementary Fig 12:**
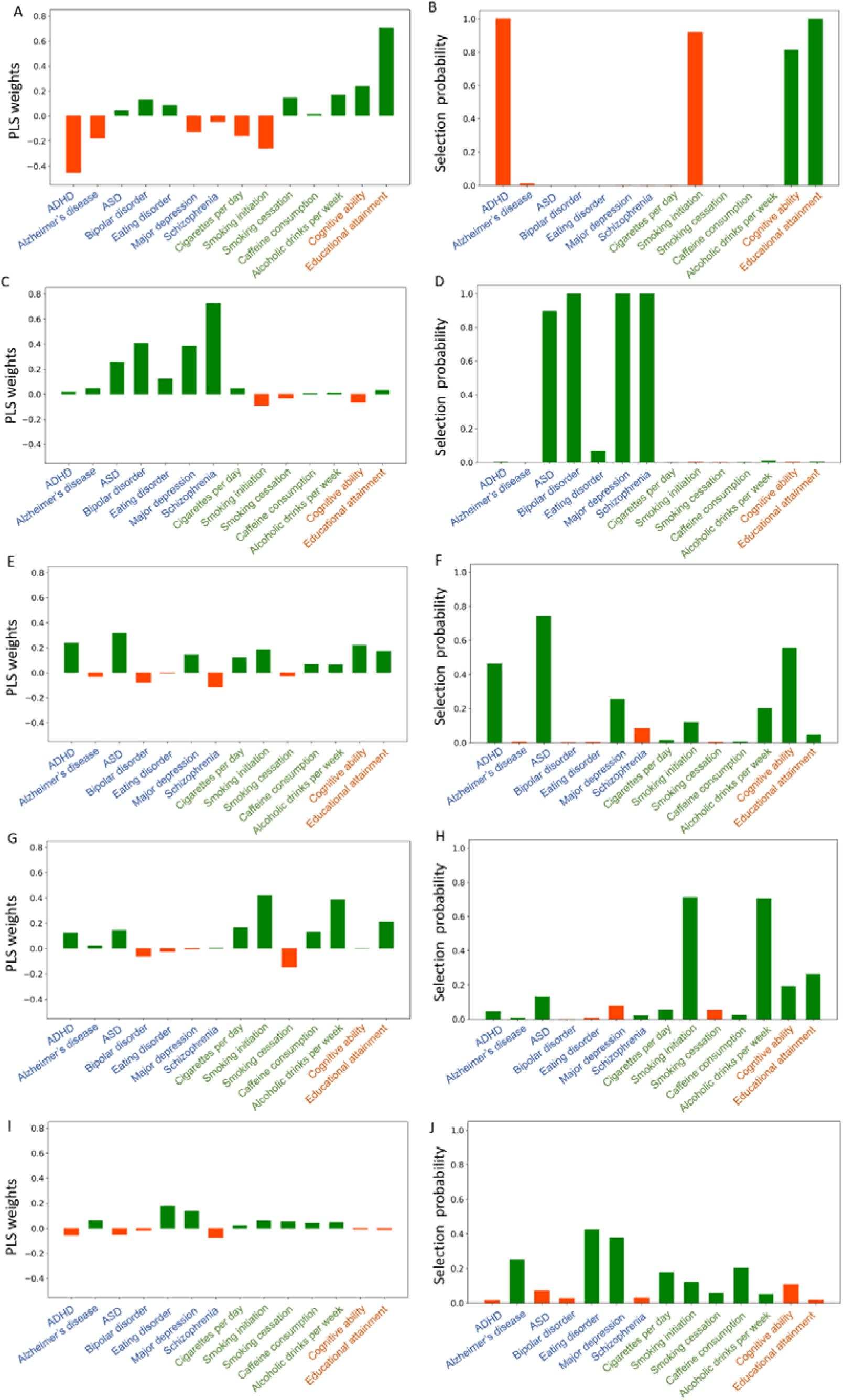
The mean weights and probabilities for first five genotype components. (A) The mean weights across 5000 repetitions for 14 PGSs in the first genotype component. (B) Probabilities of the 14 PGSs of being selected across 5000 repetitions in the first genotype component. (C) The mean weights in the second genotype component. (D) Probabilities in the second genotype component. (E) The mean weights in the third genotype component. (F) Probabilities in the third genotype component. (G) The mean weights in the fourth genotype component. (H) Probabilities in the fourth genotype component. (I) The mean weights in the fifth genotype component. (J) Probabilities in the fifth genotype component.

**Supplementary Fig.13:**
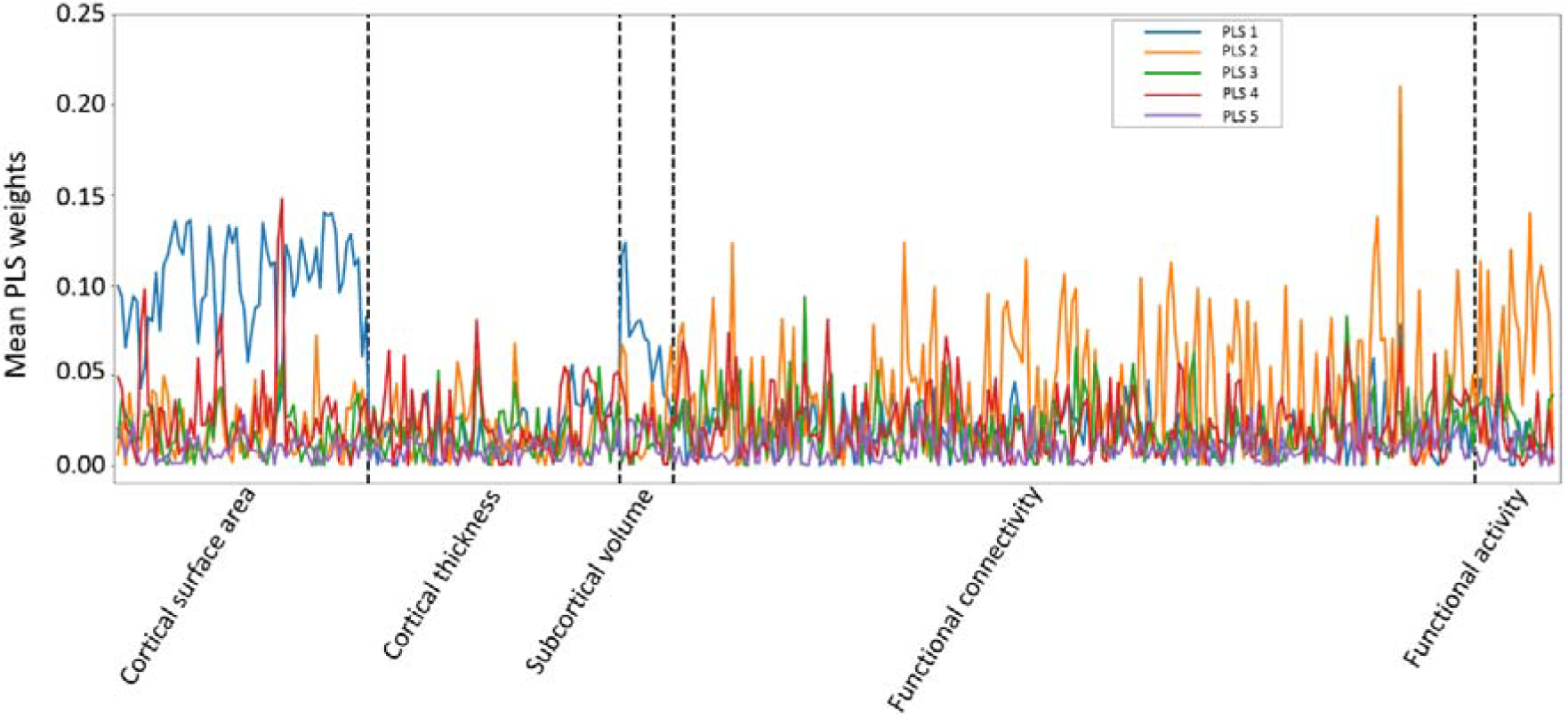
The mean weights for first five phenotype components.

**Supplementary Fig 14:**
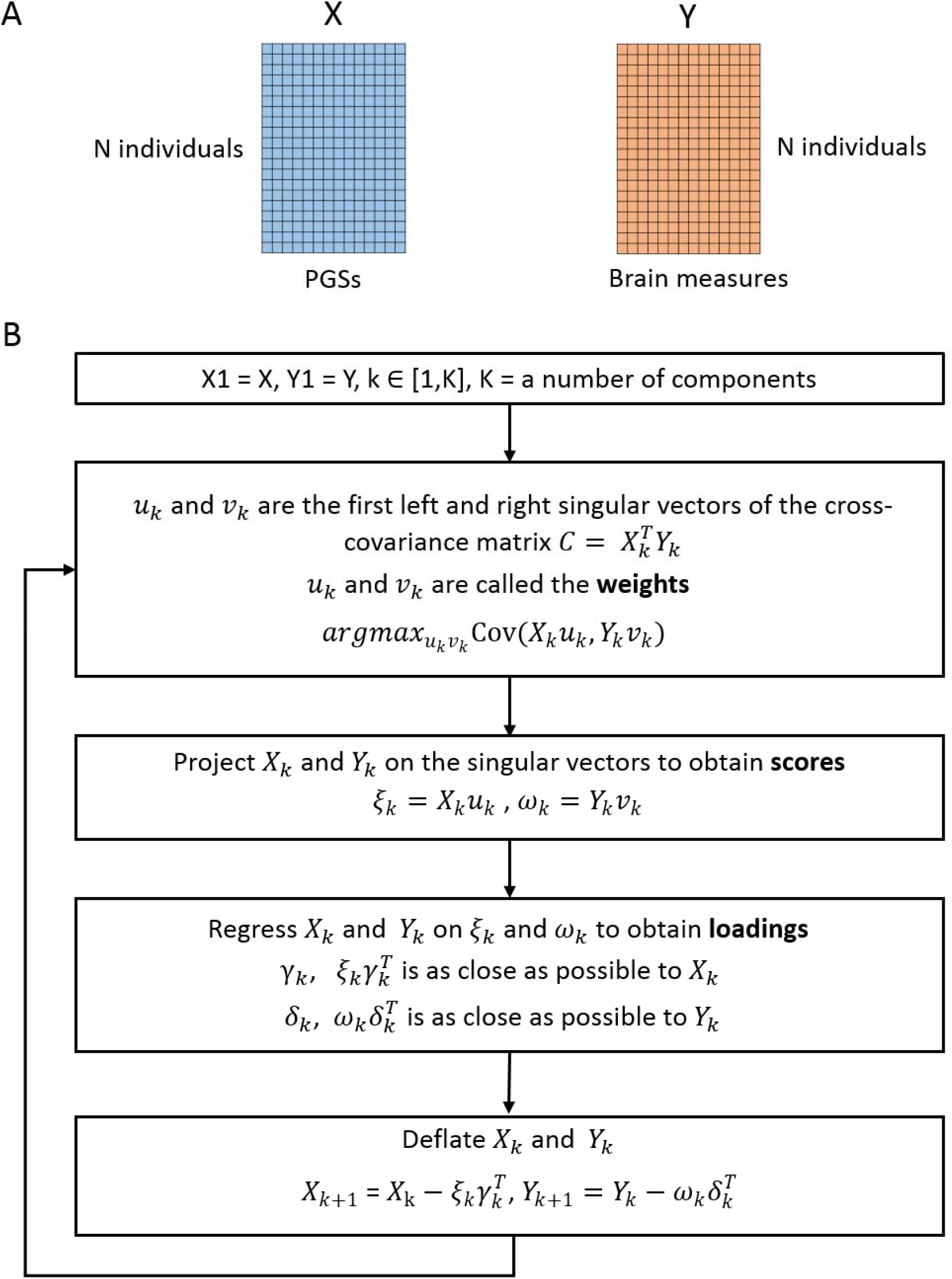
PLS procedures. (A) The input variables of PLS model. (B) The circling procedures to estimate K latent components.

## Notes

### Competing Interest Statement

The authors have declared no competing interest.

### Clinical Trial

n/a

### Author Declarations

The National Health Service North West Centre for Research Ethics Committee provided UK Biobank project with ethical approval (reference: 11/NW/0382) (http://biobank.ctsu.ox.ac.uk/crystal/field.cgi?id=200). This research has been conducted using data from UK Biobank under application numbers 40310 and 30091.

